# Time-series plasma cell-free DNA analysis reveals disease severity of COVID-19 patients

**DOI:** 10.1101/2020.06.08.20124305

**Authors:** Xinping Chen, Yu Lin, Tao Wu, Jinjin Xu, Zhichao Ma, Kun Sun, Hui Li, Yuxue Luo, Chen Zhang, Fang Chen, Jiao Wang, Tingyu Kuo, Xiaojuan Li, Chunyu Geng, Feng Lin, Chaojie Huang, Junjie Hu, Jianhua Yin, Ming Liu, Ye Tao, Jiye Zhang, Rijing Ou, Furong Xiao, Huanming Yang, Jian Wang, Xun Xu, Shengmiao Fu, Xin Jin, Hongyan Jiang, Ruoyan Chen

## Abstract

Clinical symptoms of coronavirus disease 2019 (COVID-19) range from asymptomatic to severe pneumonia and death. Detection of individuals at high risk for critical condition is crucial for control of the disease. Herein, for the first time, we profiled and analyzed plasma cell-free DNA (cfDNA) of mild and severe COVID-19 patients. We found that in comparison between mild and severe COVID-19 patients, Interleukin-37 signaling was one of the most relevant pathways; top significantly altered genes included POTEH, FAM27C, SPATA48, which were mostly expressed in prostate and testis; adrenal glands, small intestines and liver were tissues presenting most differentially expressed genes. Our data thus revealed potential tissue involvement, provided insights into mechanism on COVID-19 progression, and highlighted utility of cfDNA as a noninvasive biomarker for disease severity inspections.

**One Sentence Summary:** CfDNA analysis in COVID-19 patients reveals severity-related tissue damage.

## Main Text

A novel coronavirus, severe acute respiratory syndrome-coronavirus 2 (SARS-CoV-2) emerged at the end of 2019 (*1, 2*), resulting in outbreak of the coronavirus disease 2019 (COVID-19) across the world and more than 5 million cases were confirmed by May 25, 2020 (*3*). In a report based on ∼72,000 COVID-19 patients from China, 14% were classified as severe, 5% were critical, and the rest 81% were considered mild (*4*). Clinical progression of COVID-19 varies greatly among individuals (*4-8*), whereas the real course of the disease is not yet well understood. In fact, the incubation period for COVID-19 ranges from 1 to 14 days; the duration of viral shedding ranges from 8 to 37 days; and time from illness onset to death or discharge mainly range from 15 to 25 days (*8,9*). In addition, the case-mortality rate was found to be correlated with age, preexisting comorbid conditions such as cardiovascular disease, diabetes, and hypertension; however, reported deaths still contain high numbers of teenagers and cases without comorbidities (*4-8*). Laboratory records such as low lymphocyte counts, high C-reactive protein or D-dimer levels, and secondary bacterial infections could not provide insights into the actual process of death (*10,11*). Hence, systematical understanding of clinical course of COVID-19 and classification/prediction of severe cases precisely at early stage are essential for management of the disease.

Cell-free DNA (cfDNA) in plasma comprises short, naturally fragmented molecules that preserve valuable information related to gene expression and nucleosome footprint related to its tissues-of-origin (*12-16*). Numerous studies reported that cfDNA concentration, size profiles, coverage patterns around promoters are associated with various diseases, making cfDNA an intensively investigated biomarker for clinical use in various fields including oncology, non-invasive prenatal diagnosis, organ transplantation, autoimmune diseases, trauma, myocardial infarction, and diabetes (*13-16*). Circulating cfDNA mostly originate from died cells through apoptosis, necrosis and netosis (*14,17,18*). CfDNA is found to be potential drivers and therapeutic targets of COVID-19 suggested novel therapy strategies of the disease (14,18). However, to date, only subtotal concentration of cfDNA in serum from COVID-19 was investigated. Hence, to further explore the clinical value of cfDNA in COVID-19, we conducted systematical analysis of whole genome sequencing (WGS) data on cfDNA from mild and severe cases in time series. We report significantly different signals between mild and severe cases, which indicates potential genes, tissues, and pathways that are involved in disease course and severity, demonstrating high value in patient monitoring. Our functional analysis of cfDNA further shed light on mechanisms of progression of severe COVID-19, and demonstrated cfDNA as a potential noninvasive biomarker for disease severity inspections of COVID-19.

### Gene set enrichment analysis suggest pathways associated with severity of COVID-19

Four subjects, including two male COVID-19 patients (one mild in his 30’s and one serve in his 60’s) and 2 healthy controls (one male in his 30’s and one female in her 20’s), were recruited in this study (Table S1). For the COVID-19 patients, peripheral blood was collected at various time points within 29 days of hospitalization; plasma cfDNA was extracted and sequenced to ∼12x human haploid genome coverage for each time point (Fig. S1 and Table S2). We investigated gene expression pattern in cfDNA via analyzing the sequencing depth-normalized, relative coverage of the promoters. Gene set enrichment analysis was performed using the relative coverage around promoters of known genes on the two time-course data sets and controls (Fig. 1). Six gene clusters showing significantly different patterns between mild and severe cases were identified (Fig. 1A and Fig. 1C). Notably, the average coverage around gene promoters from cluster 2 and 6 decreased along hospitalization time line for the severe case (suggesting up-regulation of these genes), while such pattern did not exist in mild case (Fig. 1A), indicating that the genes involved in disease course could be different in mild and severe cases.

**Fig 1.**
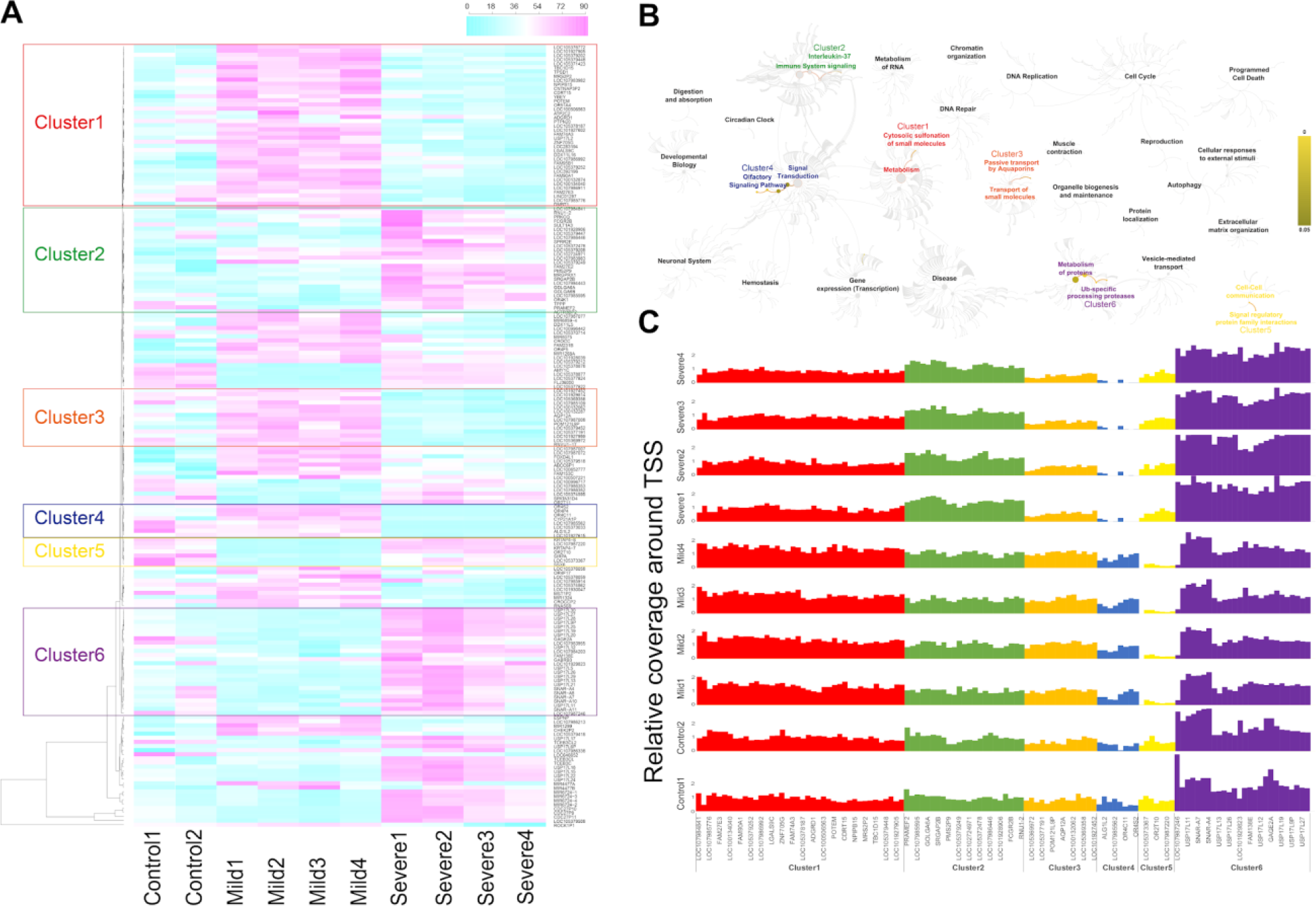
Gene set enrichment and pathway analysis on mild and severe COVID-19 cases and controls. (**A**) Heatmap on data collected at hospitalization day 11 (Mild1), 17 (Mild2), 19 (Mild3), 22 (Mild4) for mild case and day 16 (Severe1), 19 (Severe2), 25 (Severe3), 29 (Severe4) for severe case, and data from two control individuals. Color scale represents average coverage around TSSs of each gene weighted by average whole-genome sequencing depth from cfDNA. 6 clusters with differentiated pattern between mild and severe cases were marked and corresponding gene lists and relative coverage for each data set were shown (**C**). Top significant pathways based on genes of the 6 clusters were illustrated and mapped to six different functional modules (**B**).

Pathway analysis was carried out on genes of the above six clusters separately (Fig. 1B). For cluster 1 containing the maximum number of genes among the six clusters, the most enriched pathway is interleukin-37 (IL-37) signaling (*p* = 0.005, Table S3), which is involved in suppression of cytokine production and inflammation inhibition (*19, 20*).Interestingly, this pathway had been reported to have potential therapeutic effect on COVID-19 (*21*). Cluster 2 shows a clear trend of increased expression in the severe case, especially at the last three time points; significantly enriched pathways in this cluster include cytosolic sulfonation of small molecules (*p* = 0.009) and RUNX1 regulates genes involved in megakaryocyte differentiation and platelet function (*p* = 0.01), which is in accordance with the dropping platelet counts from clinical laboratory records of the severe case (Table 1). Cluster 6, which showed similar trend with cluster 2, involves Ub-specific processing proteases (*p* = 2.8×10^-9^) and deubiquitination pathways (*p* = 3.3×10^-9^), which are commonly hijacked by viruses for replication and pathogenesis, and were reported containing druggable targets to treat COVID-19 (*22*). Other enriched pathways include olfactory signaling pathway (*p* = 0.003), G alpha (s) signaling events (*p* = 0.006), signal regulatory protein family interactions (*p* = 4.5×10^-7^), and cell-cell communication (*p* = 2.3×10^-4^), which are expected given inflammatory and immune responses in severe COVID-19.

**Table 1.**
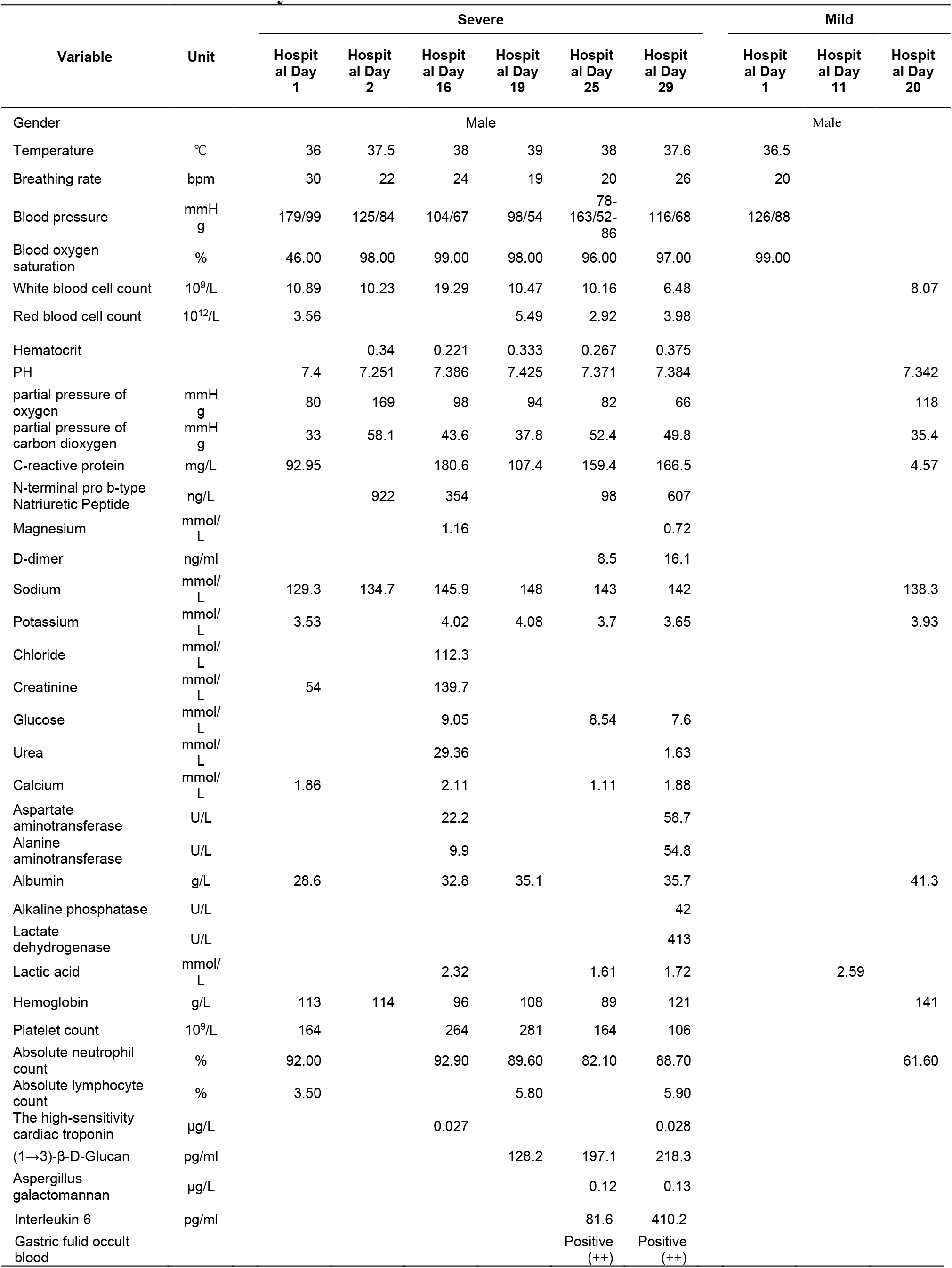
Clinical laboratory records of the mild and severe cases

### Significantly altered genes and tissue specificity on progression of COVID-19

To detect differentiated genes and tissues between mild and severe cases, relative coverages around promoters from cfDNA 1) within control group, 2) among 4 time points from mild case, 3) among 4 time points from severe case, 4) between mild and severe cases, 5) between cases and controls, were compared using Mann-Whitney rank-sum tests (Fig. 2). We laid emphasis on differences between mild and severe cases, while not significant either between cases and controls, or within controls. Table 2 lists top 10 genes out of all significantly differentially expressed genes (Fig. 2A and Table S4), and the coverage pattern around the promoters of the top 10 genes were shown in Fig. S2. Strikingly, the top 3 genes, namely POTEH (Prostate, Ovary, Testis-Expressed Protein On Chromosome 22), MGC39584 (Family With Sequence Similarity 27 Member C), and C7orf72 (Spermatogenesis Associated 48) are all highly linked to male gender: POTEH and MGC39584 are specifically expressed in prostate and testis, and C7orf72 is associated to spermatogenesis (*23*). The significant difference of genes related to male reproductive system are corroborated by previous studies that claimed spermatogonia cells to be targets attacked by SARS-CoV-2 (*24,25*). Other genes worth noting include SQLE that catalyzes oxidation of squalene, a reported conjugate of therapeutic drugs for COVID-19 (*26*), and LAMP3, which is specifically expressed in lung and associated to dendritic cell function and adaptive immunity (*27*). Additionally, the significantly differentiated expression of SPEG gene which were essential for cardiac function was consistent with clinical laboratory records (Table 1, Table S5, and Table S6) that reported the severe case with atrial fibrillation.

**Table 2.** Top 10 significantly differentiated expressed genes between mild and severe cases

| CHR | Position | Gene | Aliases of gene | p values |  |  |
| --- | --- | --- | --- | --- | --- | --- |
|  |  |  |  | Control | Case and control | Mild and severe |
| 22 | 15690025 | POTEH | Prostate, Ovary, Testis-Expressed Protein On Chromosome 22 | 0.17 | 0.24 | 2.58x10<sup>-28</sup> |
| 21 | 9821060 | MGC39584 | Family With Sequence Similarity 27 Member C | 0.08 | 1.00 | 7.44x10<sup>-27</sup> |
| 7 | 50096085 | C7orf72 | Spermatogenesis Associated 48 | 0.11 | 2.74x10<sup>-3</sup> | 2.81x10<sup>-25</sup> |
| 8 | 124999756 | SQLE | Squalene Epoxidase | 1.00 | 1.15x10<sup>-6</sup> | 1.23x10<sup>-19</sup> |
| 12 | 5043918 | KCNA5 | Potassium Voltage-Gated Channel Subfamily A Member 5 | 0.17 | 1.00 | 8.64x10<sup>-19</sup> |
| 2 | 219473003 | SPEG | Striated Muscle Enriched Protein Kinase | 0.02 | 0.04 | 1.87x10<sup>-18</sup> |
| 16 | 1773227 | EME2 | Essential Meiotic Structure-Specific Endonuclease Subunit 2 | 4.63x10<sup>-3</sup> | 0.32 | 7.22x10<sup>-18</sup> |
| 1 | 118989555 | TBX15 | T-Box Transcription Factor 15 | 0.19 | 7.96x10<sup>-3</sup> | 7.65x10<sup>-18</sup> |
| 3 | 183162878 | LAMP3 | Lysosomal Associated Membrane Protein 3 | 0.70 | 1.96x10<sup>-3</sup> | 1.23x10<sup>-17</sup> |
| 17 | 9905366 | RCVRN | Recoverin | 0.06 | 1.63x10<sup>-6</sup> | 1.83x10<sup>-17</sup> |

**Fig 2.**
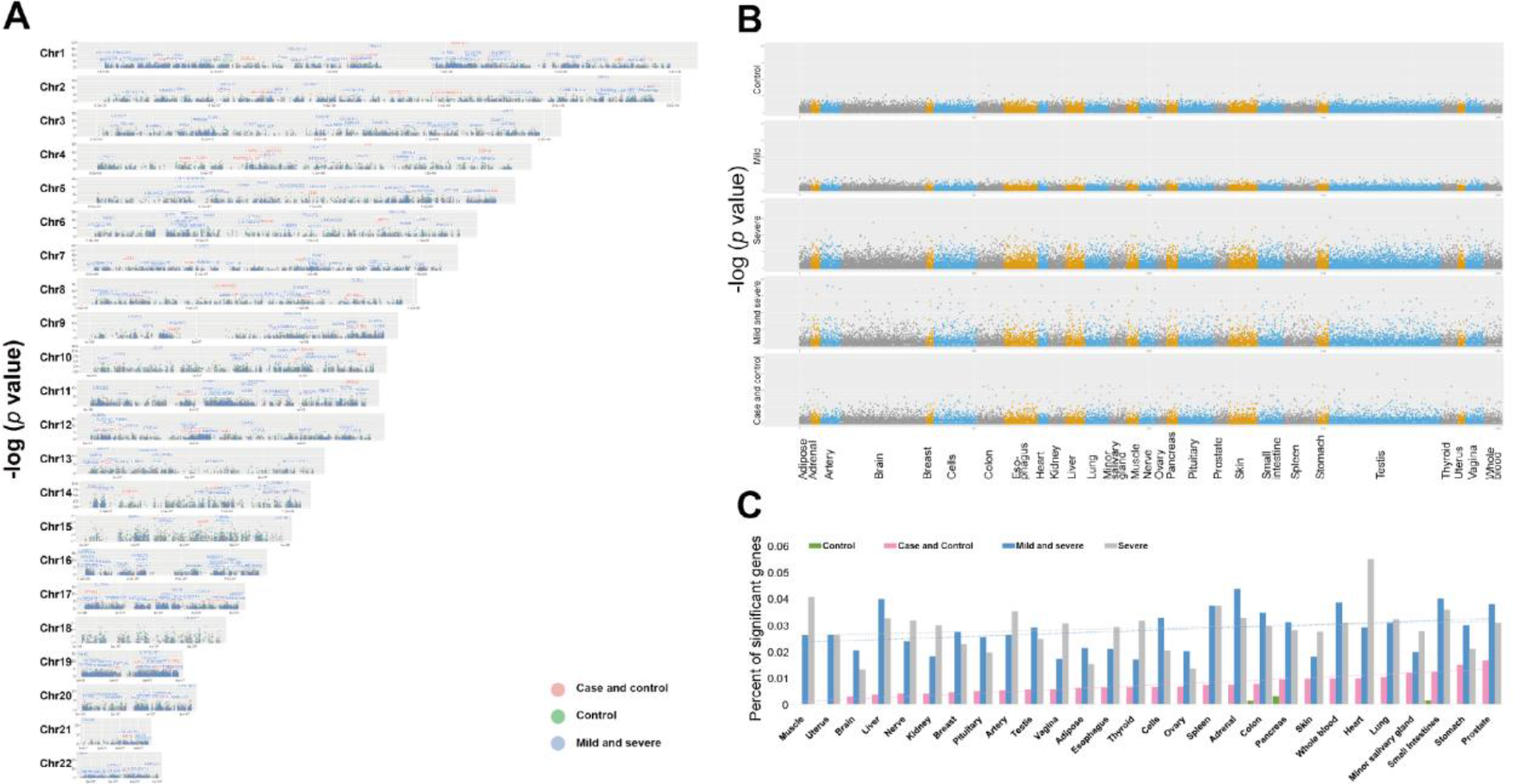
Significant tests on genes and tissues of two controls and two mild and severe cases. (**A**) Meta *p* values of rank-sum tests on coverage distribution in 200bp TSSs of genes on whole genome for case and control (pink), mild and severe (green), and control (blue). Name of genes with -log_10_(*p* value) greater than 6 within each group were marked. (**B**) Meta *p* values of genes that were expressed specifically in 28 tissues from 1) controls, 2) mild time series, 3) severe time series, 4) mild and severe cases, 5) cases and controls, from top to bottom. (**C**) Percent of significant genes marked in (**A**) within each tissue from (**B**) were summarized for control (green), case and control (pink), mild and severe (blue), severe (grey). Tissues were sorted based on percent of significant genes between case and control.

Significantly altered genes that are known to be tissue-specific (N=28) selected (Fig. 2B) and their expressing tissues were analyzed (Fig. 2C). Except for prostate and small intestines that were also significant between cases and controls, tissues that include most percent of significantly differentiated genes between mild and severe cases are adrenal, liver, and whole blood, which were acceptable in consideration of the inflammatory and immune reactions associated with COVID-19. Moreover, tissues significant specifically among data collected at the 4 time points of severe case include heart, muscle, and artery, which can be explained by the clinical records reporting arrhythmia, atrial fibrillation, and general fatigue for the severe case.

### Microbial and mitochondrial cfDNA

Besides autosomal cfDNA from cases and controls, infection of microbiomes in plasma and mitochondrial cfDNA concentration were also investigated (Fig. 3). In consistent with the RNA-virus nature of SARS-COV-2, we did not find any viral DNA of SARS-CoV-2 in the cfDNA sequencing data. Total counts of bacteria detected in plasma from COVID-19 patients were lower than that from controls (Fig. 3A), which could be explained by medication of interferons and antibiotics for these patients. Notably a novel virus infected in plasma collected at third and fourth time points of the severe case was human betaherpesvirus 5 (Table S7), which might cause pneumonia, colitis, or encephalitis in immunocompromised people (*28*).

**Fig 3.**
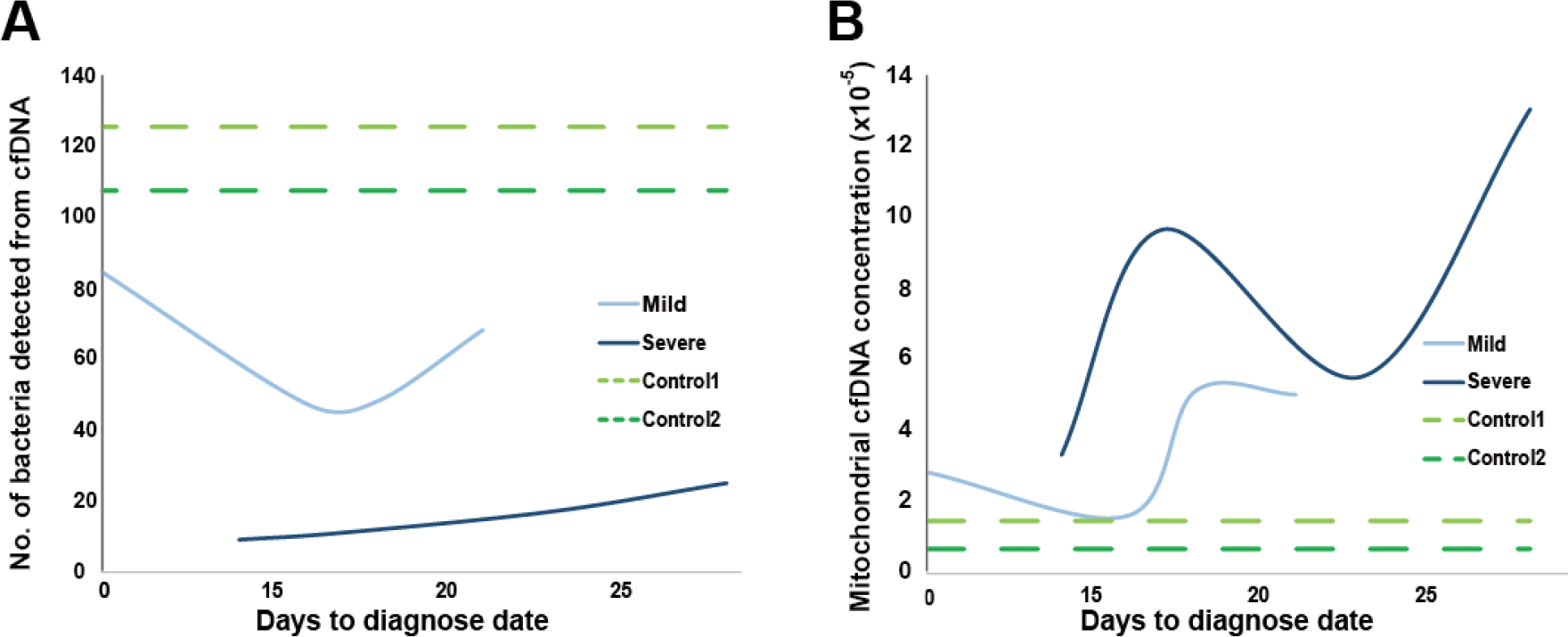
Microbial and mitochondrial cfDNA from two controls and two mild and severe cases. No. of bacteria detected (**A**) and mitochondrial cfDNA concentration (**B**) from cfDNA of plasma collected at 4 time points for mild (light blue) and severe (dark blue) cases, and at once for the two cases (green, time for dotted lines are invented only for comparison).

Overall, mitochondrial cfDNA concentrations of plasma from cases were lower than controls, while the severe case had higher concentration than mild case (Fig. 3B). Notably distribution of mitochondrial concentration for severe case showed clear “S” shape, which was matched with hematocrit and hemoglobin at last four collection time (Table 1), suggesting hypoxia of the patient.

## Discussion

Acute progression of COVID-19 makes it vital to discriminate patients at high risk to go through rapid deterioration. Results of this study on cfDNA from patients with COVID-19 revealed signals associated with disease severity. We found that IL-37 signaling pathway and SQLE gene are both altered in COVID-19 patients, which results were consistent with previous reports. We also discovered signals that are in line with clinical records, including differentially expressed genes related to atrial fibrillation and male reproduction system. In addition, we observed changes of mitochondrial cfDNA concentration, which matches with hematocrit and hemoglobin of the patient. Moreover, alterations in Ub-specific processing proteases and deubiquitination pathways, SQLE, LAMP3, and SPEG genes provide clues on drug targets and biomarkers for COVID-19. Considering the relatively small sample size, follow-up analyses with large-scale sample size is required to make solid conclusions. Nevertheless, the preliminary results demonstrate that cfDNA could serve as a valuable analyte for surveillance, medication guidance, and prognosis of COVID-19 patients.

## Data Availability

The data that support the findings of this study have been deposited into CNGB Nucleotide Sequence Archive (https://db.cngb.org/cnsa/) with accession number CNP0001081, and the availability of the data will be confirmed as soon as approval of the study is signed by Chinese human genetic resource committee.

## Acknowledgments

This work was supported by China National GeneBank. We sincerely thank the support provided by China National GeneBank.

## Author contributions

Ruoyan Chen, Xin Jin, Hongyan Jiang, and Shengmiao Fu had full access to all of the data in the study and take responsibility for the integrity of the data and the accuracy of the data analysis; concept and design: Xin Jin, Xinping Chen, Ruoyan Chen; acquisition, analysis, or interpretation of data: Ruoyan Chen, Yu Lin, Jinjin Xu, Yuxue Luo, Tingyu Kuo, Ye Tao; drafting of the manuscript: Ruoyan Chen, Kun Sun; critical revision of the manuscript for important intellectual content: Xin Jin, Xun Xu, Kun Sun; statistical analysis: Ruoyan Chen, Yu Lin, Jinjin Xu; obtained funding: Xun Xu, Xin Jin, Xinping Chen, Huanming Yang, Jian Wang; administrative, technical, or material support: Tao Wu, Zhichao Ma, Hui Li, Chen Zhang, Jiao Wang, Xiaojuan Li, Feng Lin, Junjie Hu, Ming Liu, Jiye Zhang, Fang Chen, Chunyu Geng, Chaojie Huang, Jianhua Yin, Rijing Ou, Xiaorong Fu; supervision: Xin Jin, Xinping Chen, Hongyan Jiang, Shengmiao Fu.

## Data and materials availability

The data that support the findings of this study have been deposited into CNSA (CNGB Nucleotide Sequence Archive) of CNGBdb with accession number CNP0001081 (https://db.cngb.org/cnsa/).

## Supplementary Materials for

### Materials and Methods

#### Data collection

A total of 10 samples were collected from 2 patients with COVID-19 at 4 time points and 2 health controls. Leftover surplus blood was collected from participants who had signed consent forms after clinical diagnosis. Peripheral blood was stored using EDTA anticoagulant-coated tubes. The blood samples pretreatment and DNA extraction were proceeded at a Biosafety Level 2 (BSL-2) laboratory to ensure the appropriate biosafety practices (*1*). All samples were centrifuged at low speed (3000 rpm) for 10 min at 4°C within six hours after collection. The supernatant was centrifuged at high speed (1,4000 rpm) for 10 min at 4°C. Then the plasma was set at 56°C water bath for 30 minutes. Circulating cfDNA was extracted from 200uL plasma using MagPure Circulating DNA Mini KF Kit (MD5432-02) following the manufacturer’s guide. The cfDNA was eluted by 200uL TE buffer for QC and 40uL for the rest. For cfDNA library construction, the extracted cfDNAs were processed to library using MGIEasy Cell-free DNA Library Prep kit (MGI, cat. No.: AA00226).

For upstream data processing, firstly, soapnuke (version 1.5.0) (*2*) was used for trimming the sequencing adapters from raw reads, and filtering low quality and high ratio N. N rate threshold was set to 0.1, low quality rate was set to 0.5, and low quality threshold, namely the max mismatch number when match the adapter to 2 and the clean data quality system to sanger, was set to 12. Secondly, BWA (version 0.7.17-r1188) (*3*) was used to map reads against the human reference genome (build hg38). After sorting and removing duplication of the aligned reads, variants were detected by Haplotype caller from GATK (*4*). The above steps were performed by sention (*5*), a platform for processing of genomic data that combined alignment, variants calling, and quality recalibration together efficiently.

#### Gene set enrichment and pathway analysis

Firstly, for each gene, average sequencing depth within 2kb region around transcription start sites (2kb-TSSs) and 200bp region around TSSs (200bp-TSSs) were calculated respectively by DepthOfCoverage package from GATK (*4*). The relative coverage around TSS was the above depth normalized by average depth of WGS data from each sample. Secondly, for each gene, S_i_ = D_maxi_ - D_mini_ was calculated, where D_maxi_ represented the highest depth among group *i* within 2kb-TSSs/200bp-TSSs and D_mini_ represented the lowest depth among group *i* within 2kb-TSSs/200bp-TSSs. Genes with S_controls_ > S_cases_ were filtered, and remained genes were ranked by S_cases_. Gene set enrichment analysis was performed on the top 1% ranked genes by the heatmap package from R version 3.5.1, and clusters of genes were selected from dendrogram output by heatmap package. Pathway analysis was carried out by Reactom (*6*) on the 6 clusters of genes separately (Table S2). Results based on 2kb-TSS regions were presented in Fig. 1, and results based on 200bp-TSS regions were summarized in Fig. S3.

#### Analysis on differentiated genes and tissue specificity

To test difference among different groups, including 1) G_control_: 2 samples from 2 control individuals, 2) G_mild_: 4 samples from 1 mild case at 4 time points, 3) G_severe_: 4 samples from 1 severe case at 4 time points, 4)G_mild,severe_: 4 mild and 4 severe samples, 5) G_case,control_: 2 controls and 8 cases, reads start counts, namely counts of starts of sequencing reads pairs, were calculated within 200bp region around transcription start site (TSS) of each gene. For the 2 samples from 2 control individuals and the 8 samples from 4 cases collected at 4 time points, pairwise Wilcox’s Rank-sum tests were carried out by wilcox package from R version 3.5.1, and corresponding *p* values, including G_control_∈{*p*_control1,control2_}, G_mild_∈{*p*_mild1,mild2_, *p*_mild1_mild3_, *p*_mild1_mild4_, *p*_mild2_mild3_, *p*_mild2,mild4_, *p*_mild3_mild4_}, G_severe_ ∈{*p*_severe1,severe2_, *p*_severe1,severe3_, *p*_severe1,severe4_, *p*_severe2,severe3_, *p*_severe2,severe4_, *p*_severe3,severe4_}, G_mild,severe_∈{*p*_mild1,severe1_, *p*_mild1,severe2_, *p*_mild1,severe3_, *p*_mild1,severe4_, *p*_mild2,severe1_, *p*_mild2,severe2_, *p*_mild2,severe3_, *p*_mild2,severe4_, *p*_mild3,severe1_, *p*_mild3,severe2_, *p*_mild3,severe3_, *p*_mild3,severe4_, *p*_mild4,severe1_, *p*_mild4,severe2_, *p*_mild4,severe3_, *p*_mild4,severe4_}, G_case,control_∈{*p*_control1,mild1_, *p*_control1,mild2_, *p*_control1,mild3_, *p*_control1,mild4_, *p*_control1,severe1_, *p*_control1,severe2_, *p*_control1,severe3_, *p*_control1,severe4_, *p*_control2,mild1_, *p*_control2,mild2_, *p*_control2,mild3_, *p*_control2,mild4_, *p*_control2,severe1_, *p*_control2,severe2_, *p*_control2,severe3_, *p*_control2,severe4_,}, were generated. For each group of *p* values above, a combined *p* value was calculated using Wilkinson’s method (*7*). Cutoff for selection of significantly differentially expressed genes of each group was set to 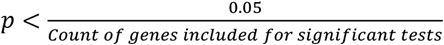, namely, *p* < 1.6 × 10^−6^.

Tissue specificity was analyzed based on grouped meta *p* values above. 23,570 transcripts that are expressed specifically from 28 tissues were selected based on GTEx database (*8,9*).

#### Microbiomes and mitochondrial cfDNA concentration estimation

The sequences remaining after removal of reads identified as human were aligned to microbial genome databases, contains 4153 viral and 2328 bacterial genomes, 208 fungi and 79 parasites genomes, coronavirus 2 (SARS-CoV-2) genome downloaded from NCBI, NC_045512.2. For identifying a positive microbe, we used the methodological criteria according to (*10-12*), focused on a). Coverage rate; b). Species level sequencing depth; c). Species level relative abundance and d). Unique mapped reads numbers.

Concentration of mitochondrial cfDNA was calculated as count of cfDNA fragments (sequencing reads pairs) that were uniquely mapped to mitochondrial DNA divided by total count of cfDNA fragments that were uniquely mapped to whole genome (Build hg38).

**Fig S1.**
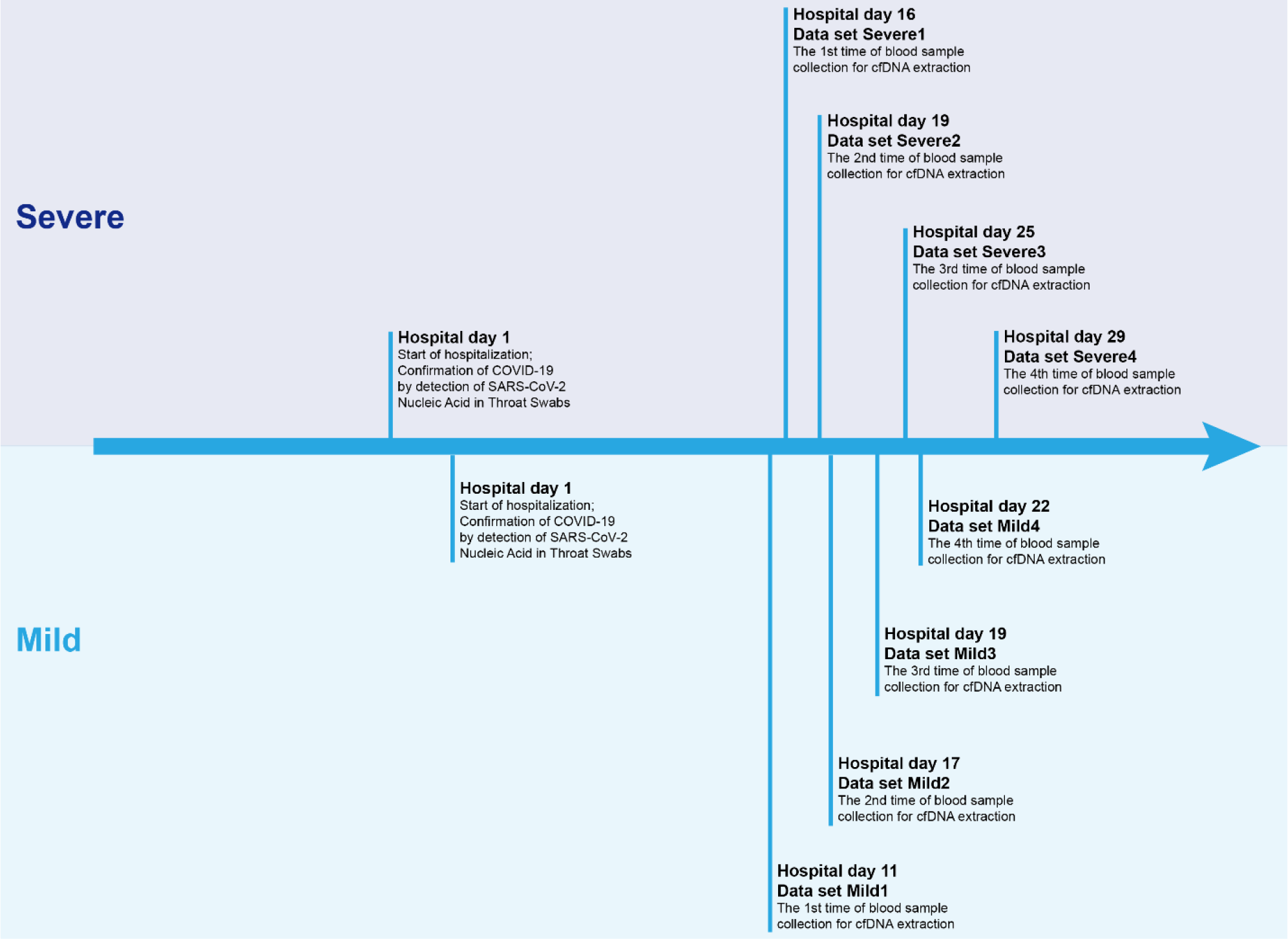
Illustration of timelines for the mild and severe cases.

**Fig S2.**
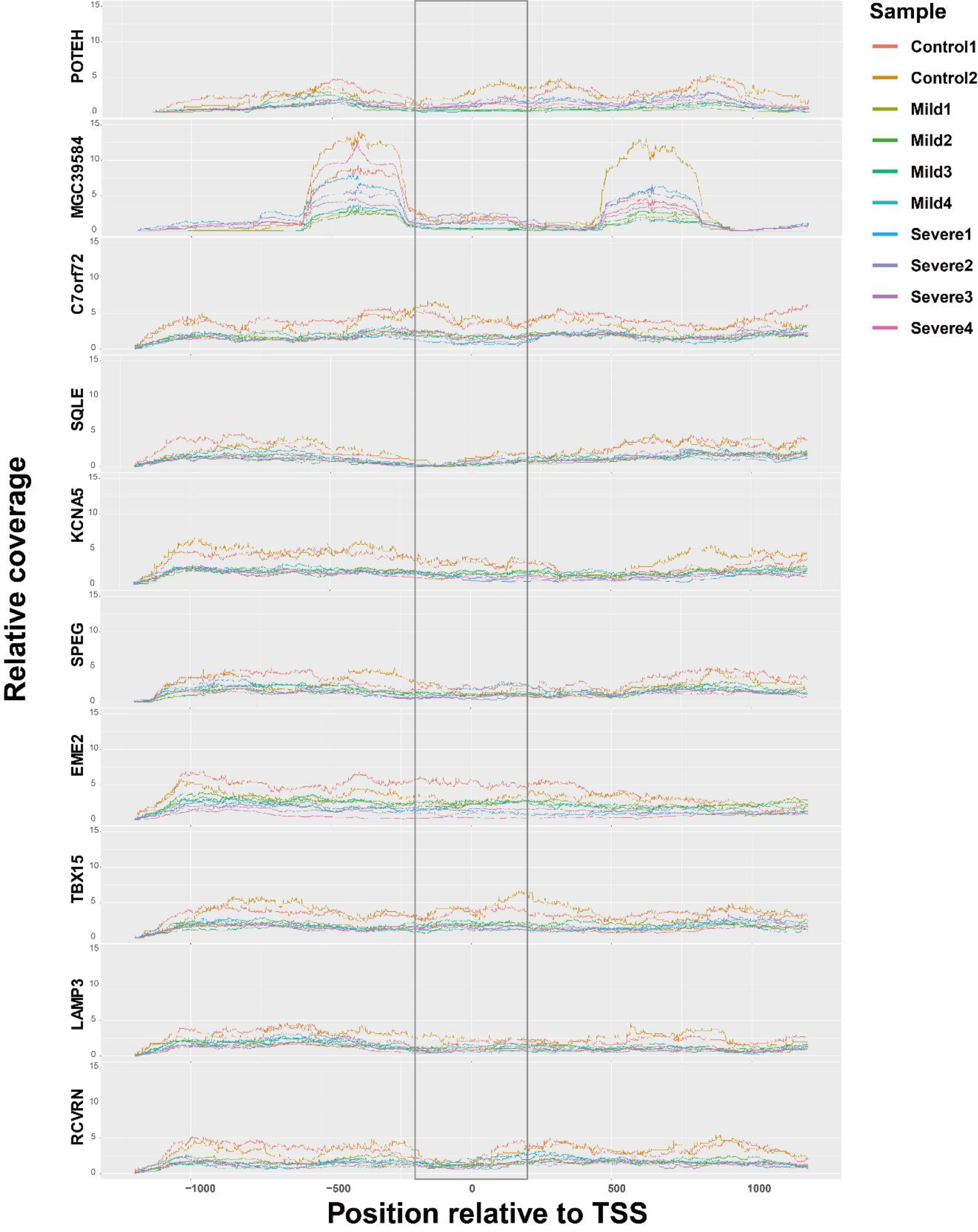
Distribution of coverage around TSSs of genes significantly different between severe and mild cases. Relative coverage was calculated as coverage of cfDNA fragments weighted by average sequencing depth of each sample. Top 10 significant genes were ranked by *p* values from top to bottom. Regions within the grey rectangle were used for significant test.

**Fig S3.**
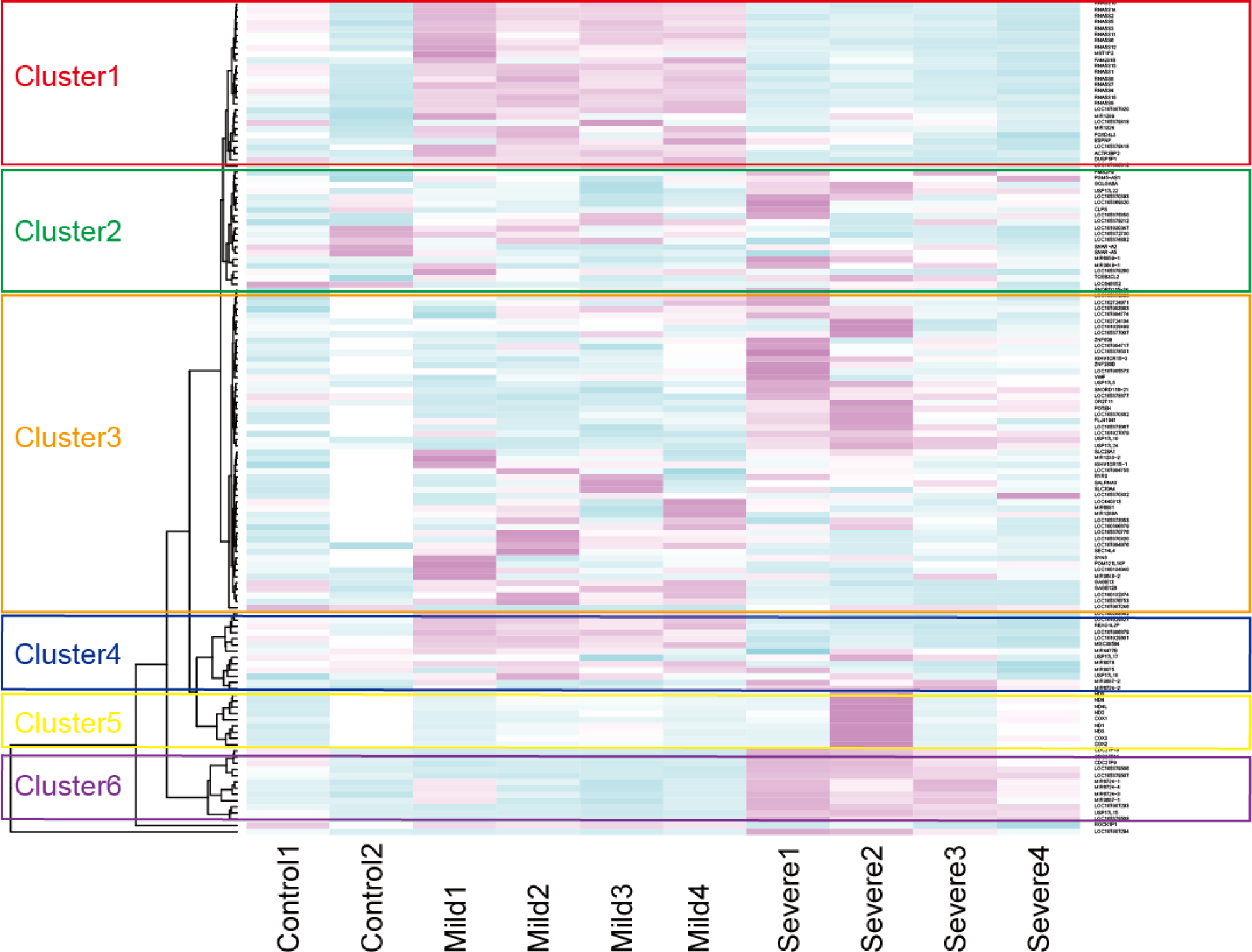
Gene set enrichment analysis based on relative coverage around 200bp-TSSs. Same gene set enrichment analysis as Fig. 1 was performed on 200bp-TSSs regions alternatively, and 6 clusters of genes were used for pathway analysis (Table S8).

**Table S1.**
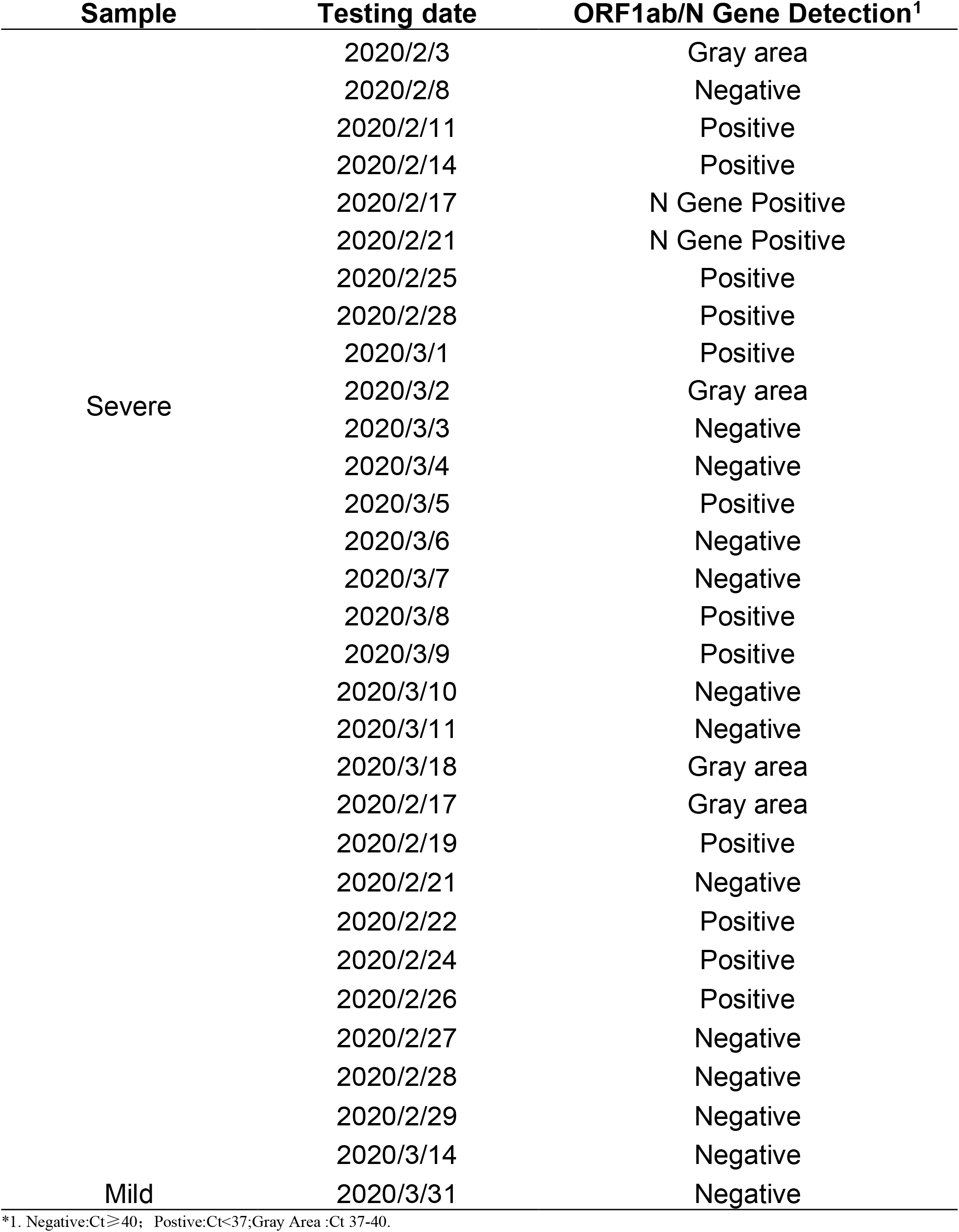
Summary on PCR testing of throat swab nucleic acids results.

**Table S2.**
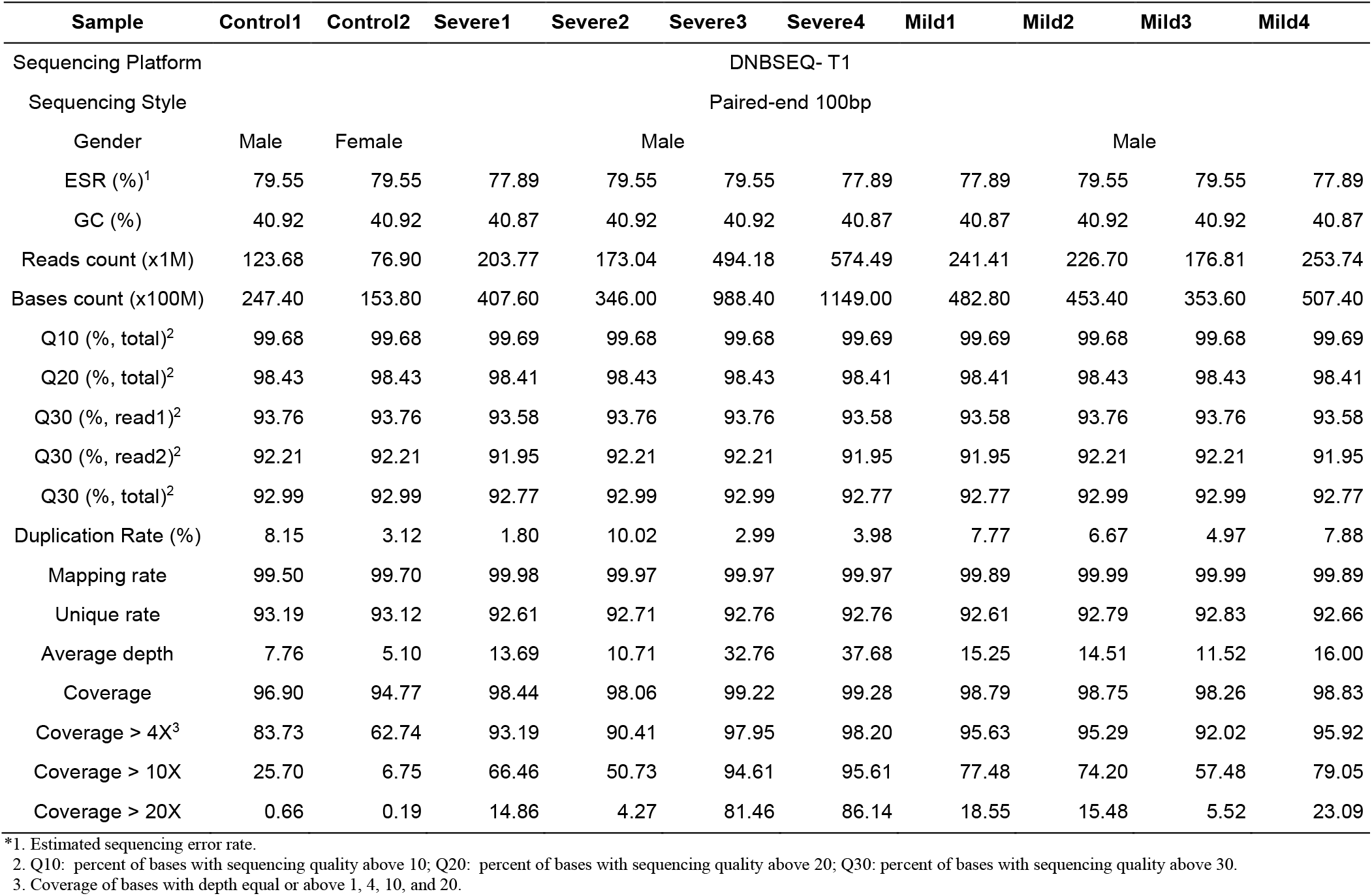
Summary on WGS data of the 2 cases and 2 controls.

**Table S3.**
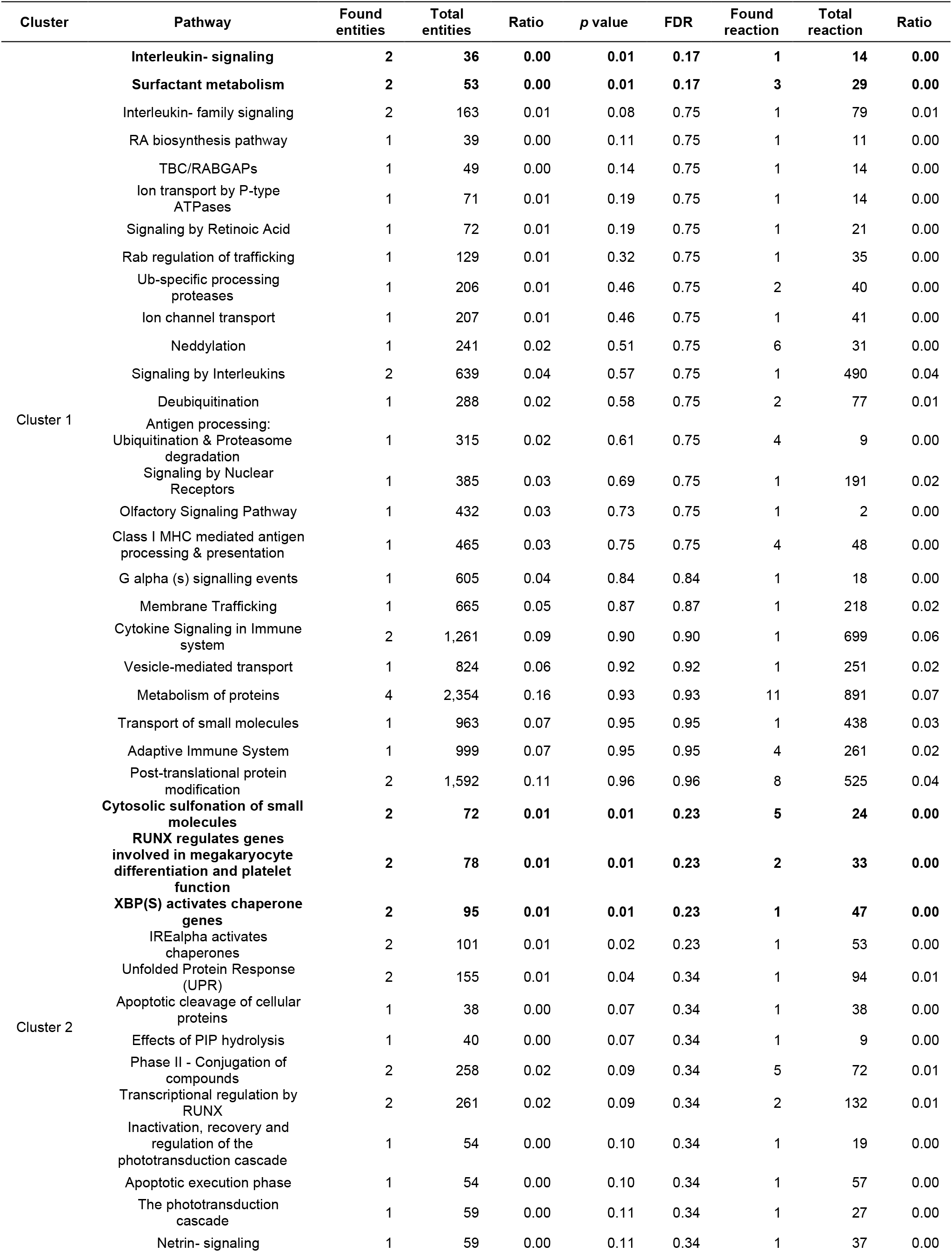
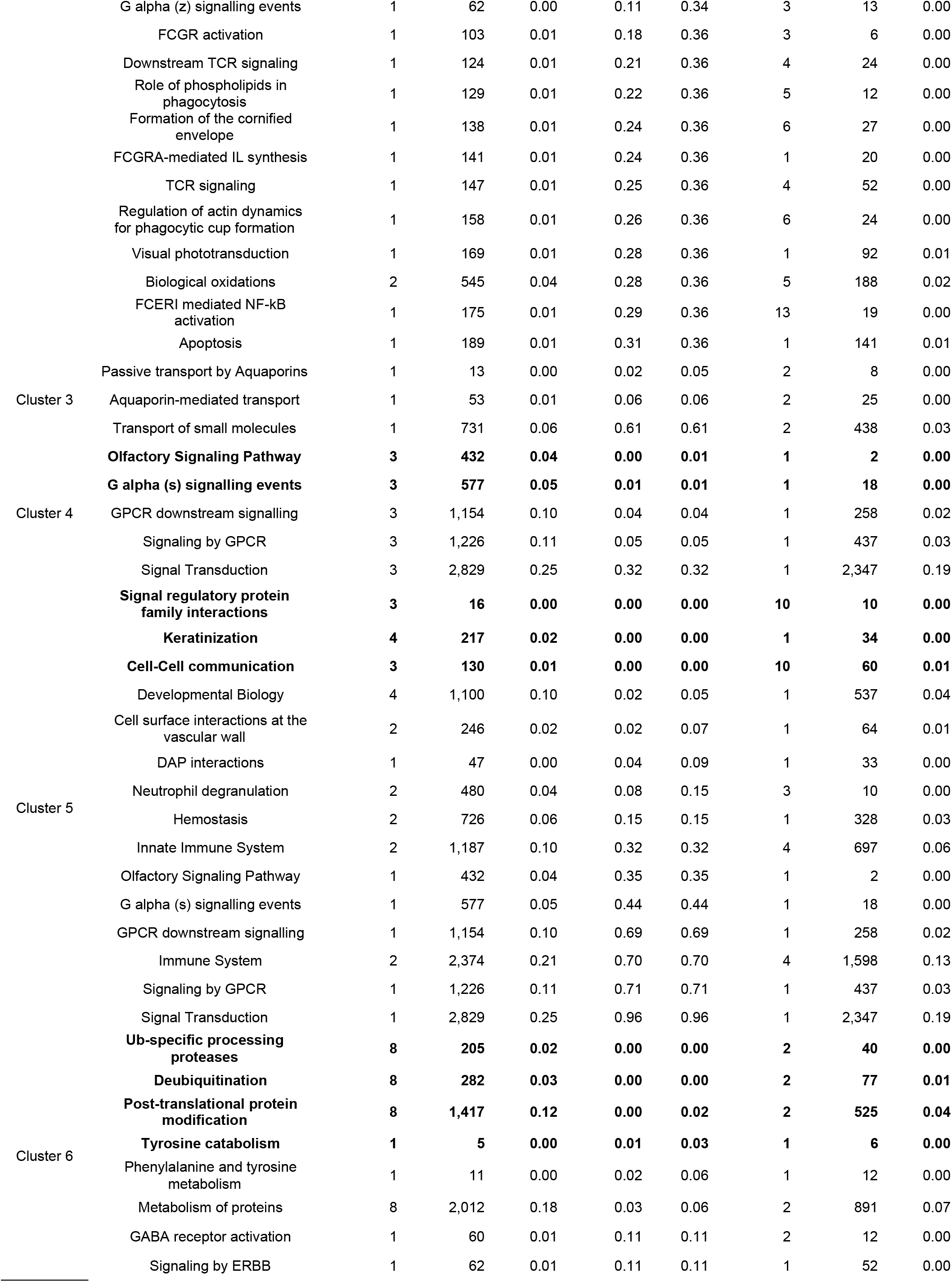
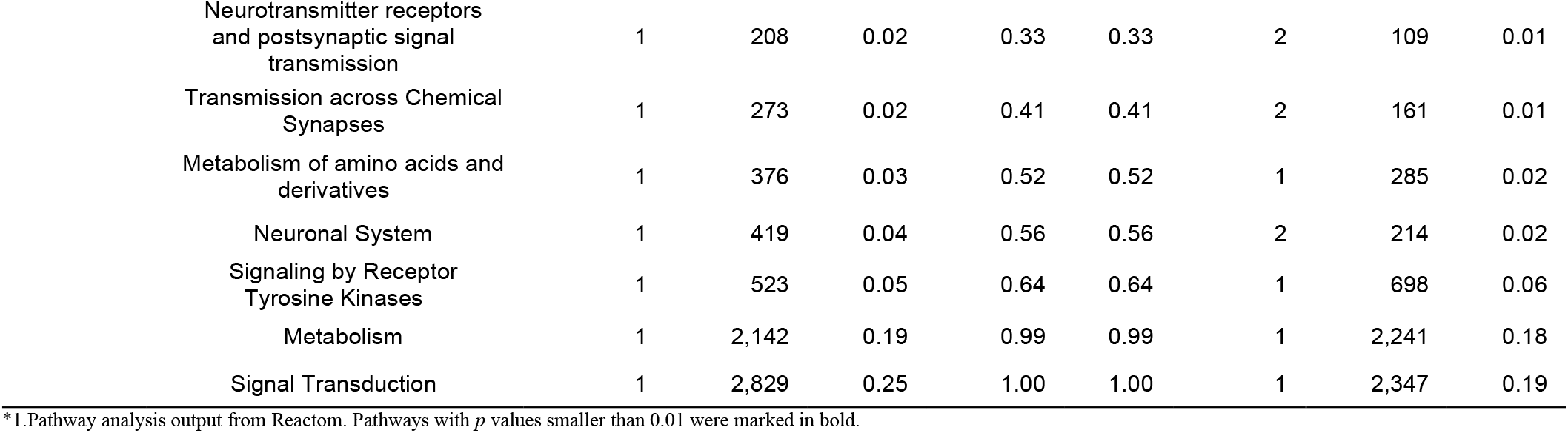
Results on pathway analysis of the 6 clusters from coverage around 2kb-TSSs from cfDNA of one mild case, one severe case, and two controls^1^.

**Table S4.**
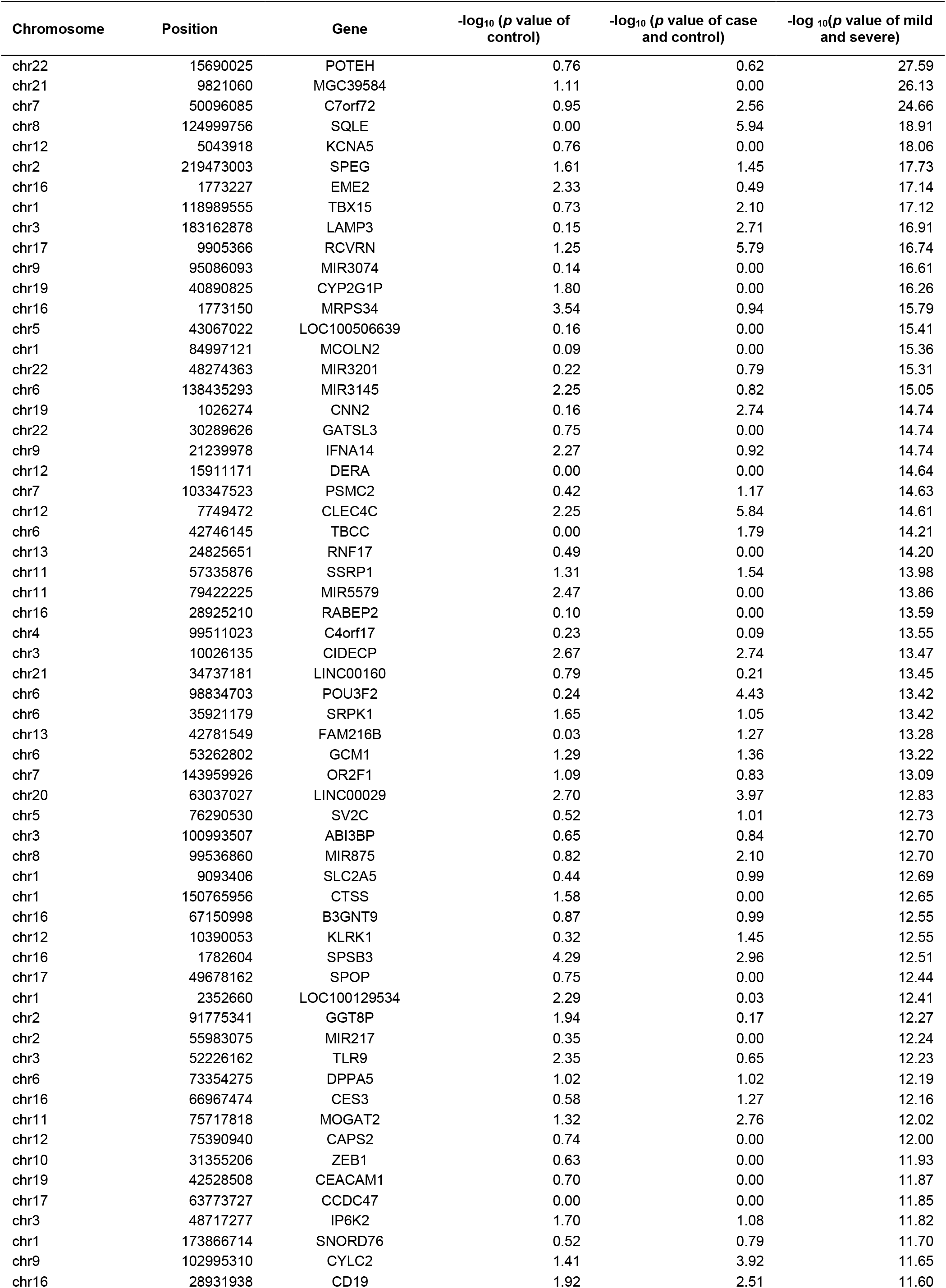
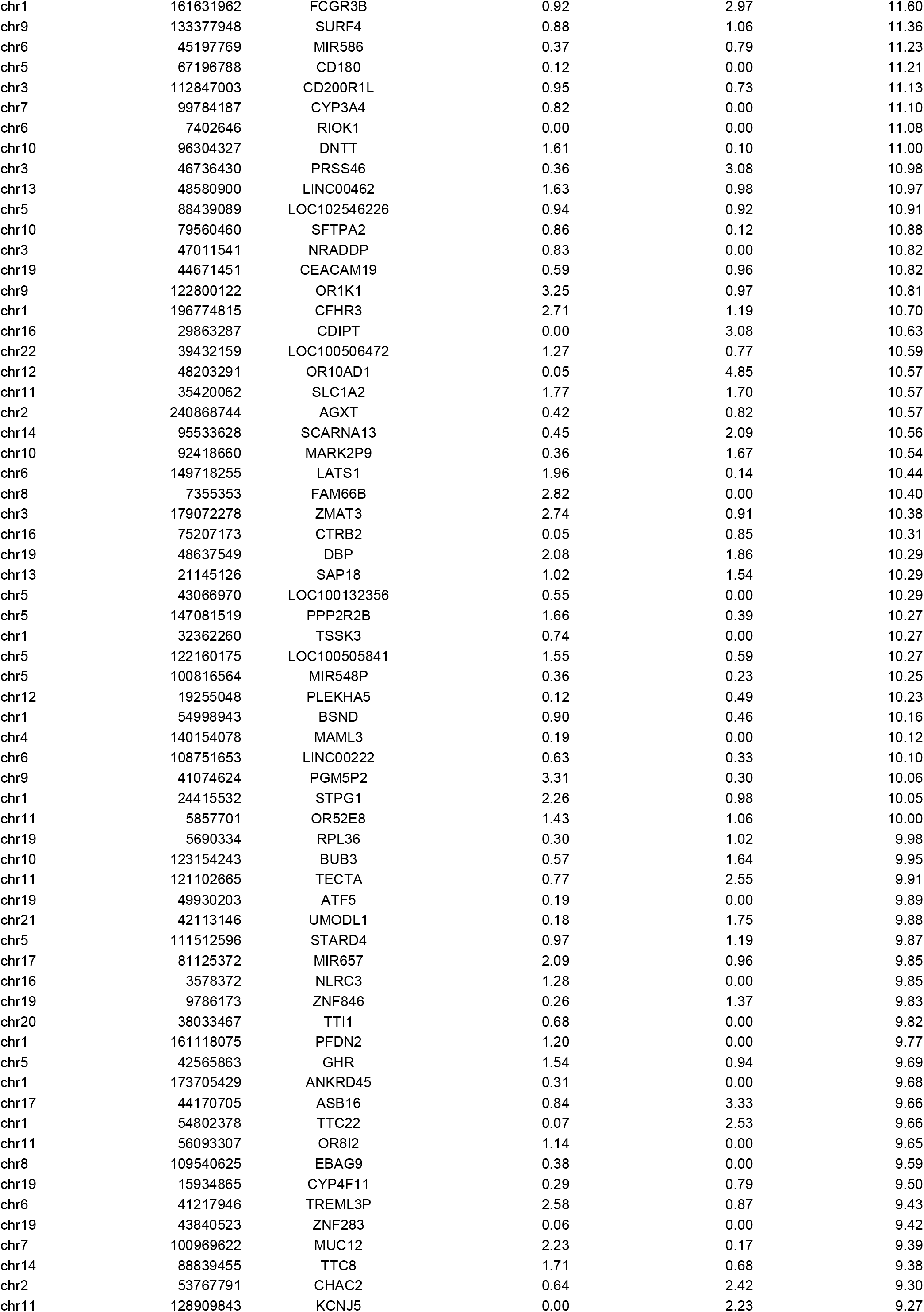
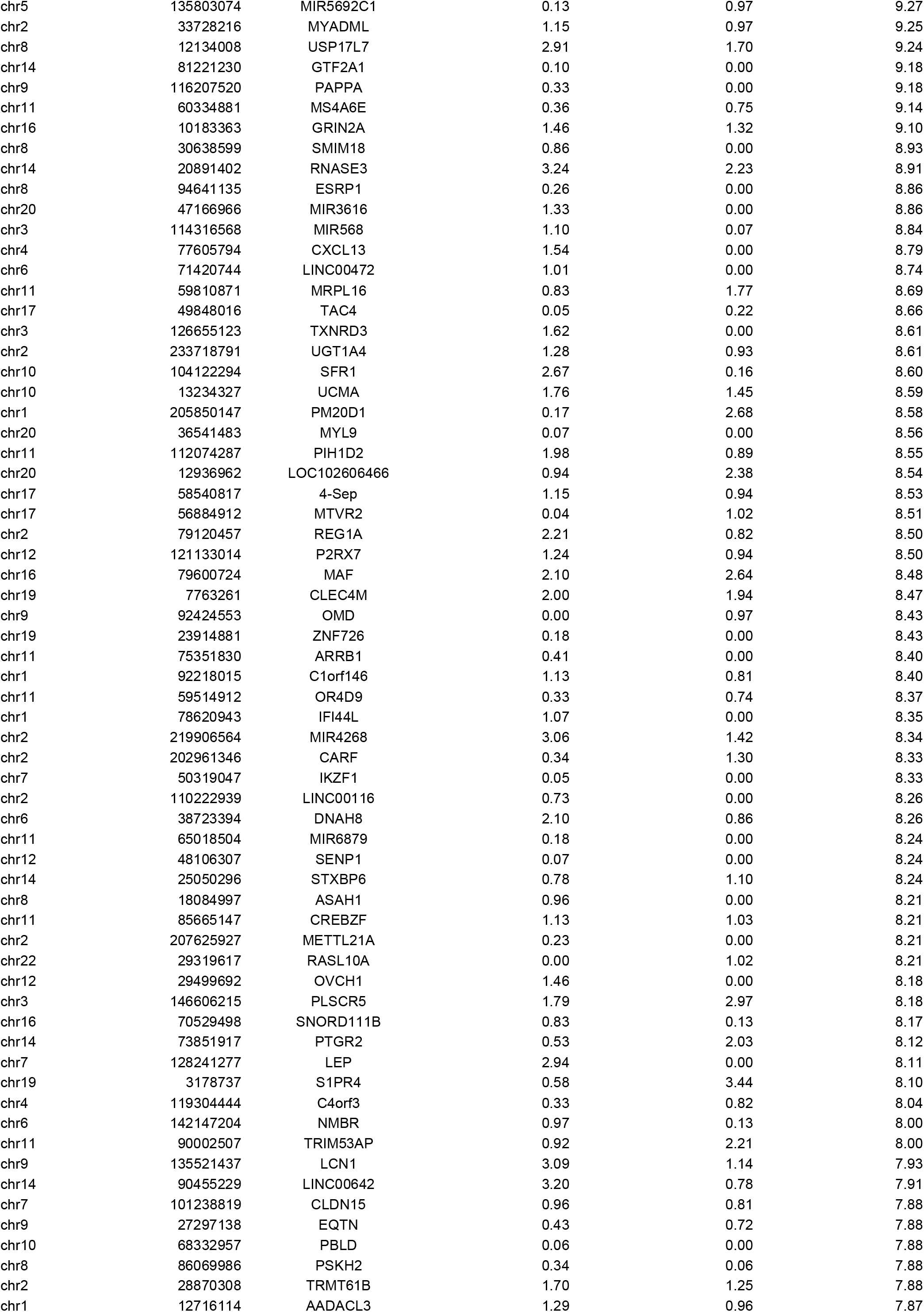
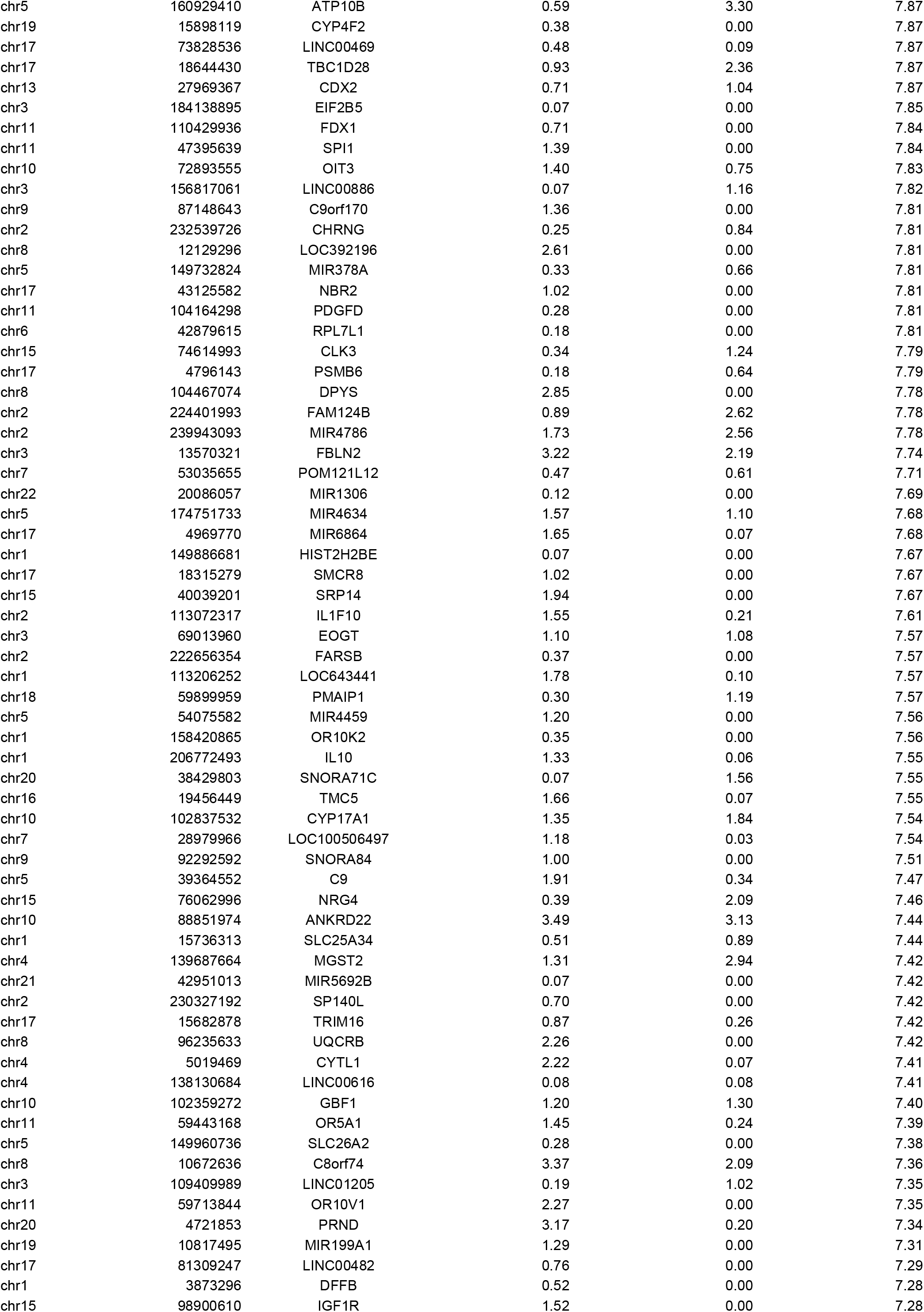
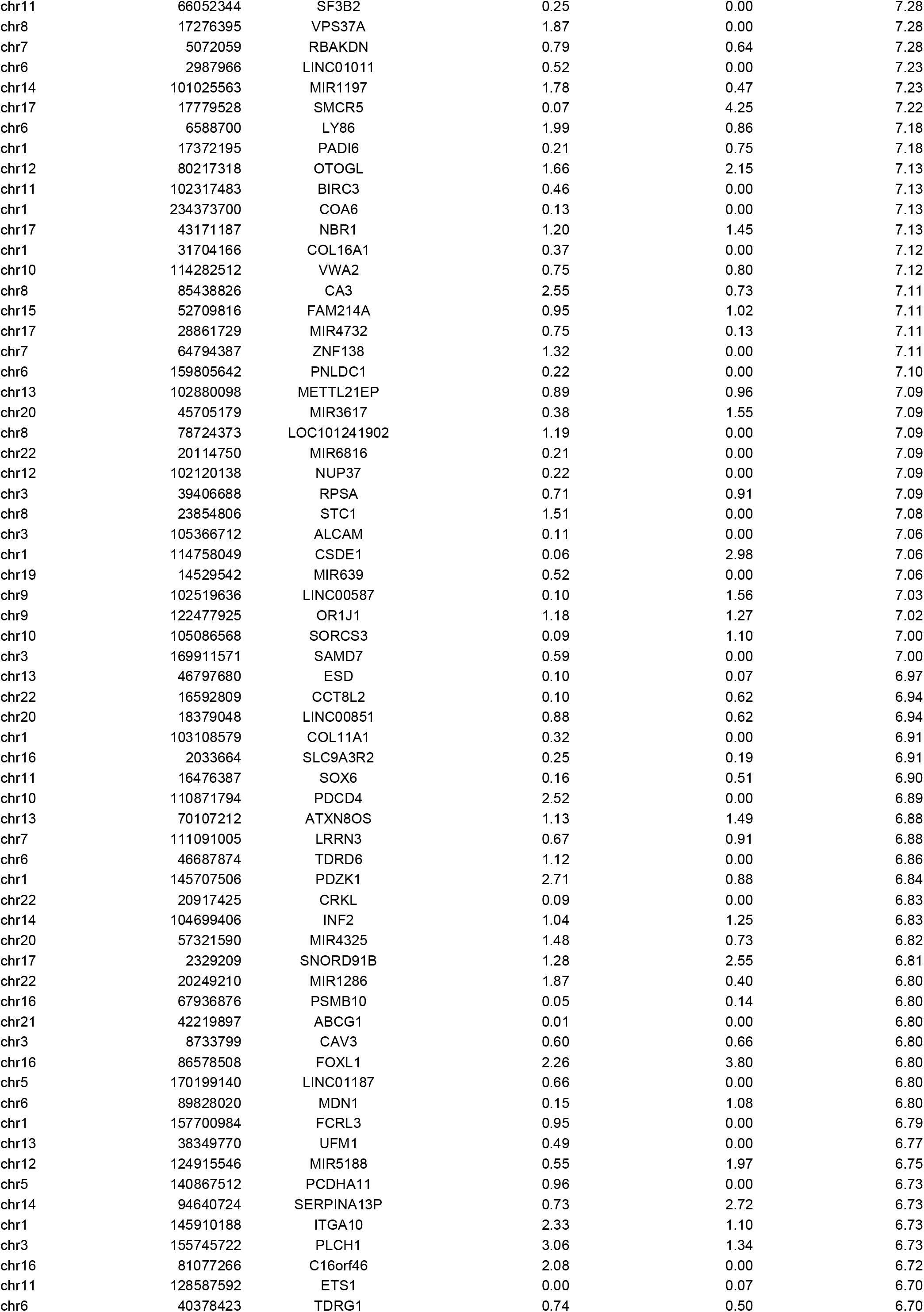
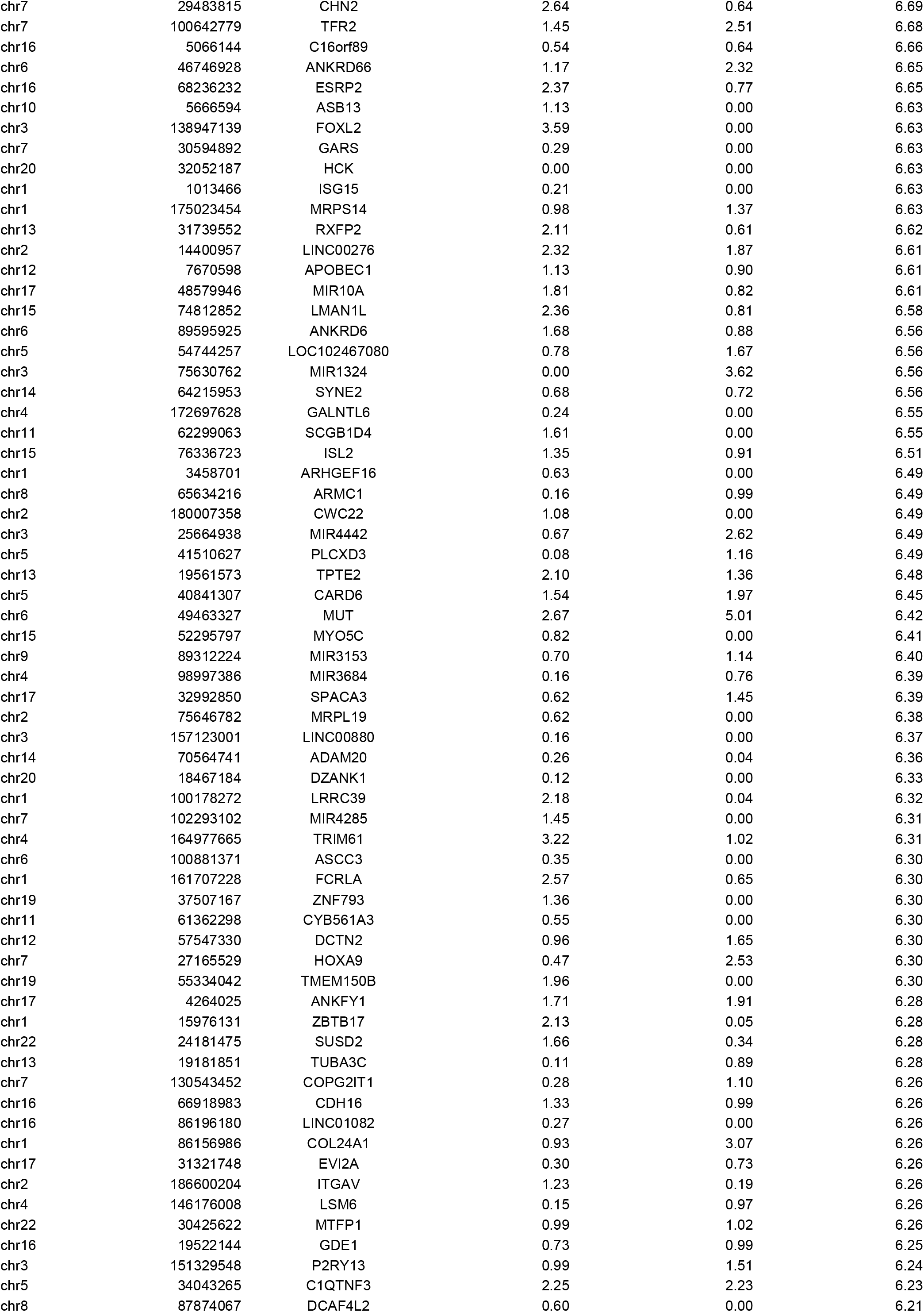
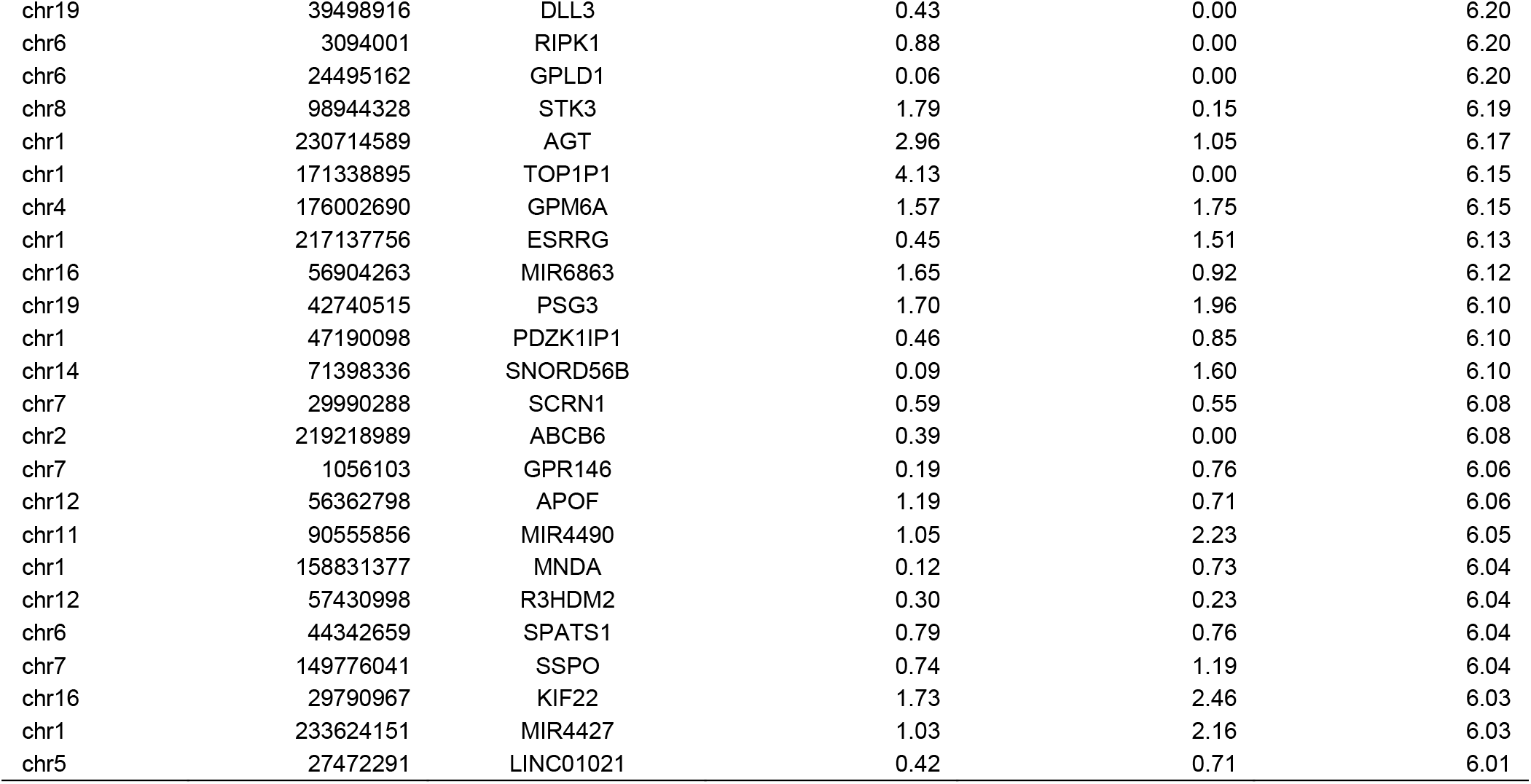
All significantly different genes between mild and severe cases^1^.

**Table S5.** Clinical medication records of the severe case.

| Date | Clinical medication | Increase/Decrease Dosage | Auxiliary Survival |
| --- | --- | --- | --- |
| Hospital Day 2 | Use Doxycycline<br>Use Interferon atomization<br>Use Methylprednisolone (80mg)<br>Use Thymopentin + Immunoglobulin (10g)<br>Use Meropenem + Linezolid<br>Use Abidor + Kridge<br>Use Xuebijing + Ulinastatin<br>Use Norepinephrine<br>Use Mosapride<br>Use Human albumin (10g) |  | Invasive Ventilator ( SIMV mode<br>VT 450 mL<br>PS 15 cmH2O<br>PEEP 12 cmH2O<br>FiO2 70%<br>Respiration ratio 1:1.5<br>Frequency 20 bpm.<br>)<br>Right subclavian vein puncture ca<br>theterization<br>Right femoral vein catheterization |
| Hospital Day 3 | Stop Linezolid<br>Stop Kridge<br>Use Midazolam<br>Use Morphine + Fentanyl<br>Use Amiodarone<br>(Intravenous injection)<br>Use Enteral nutrition<br>(1000 mL) | Maintain CRRT 12 hr.<br>Maintain Meropenem<br>Maintain Thymopentin | Invasive Ventilator ( Decrease PS from 15 to 12 cm H2O<br>Decrease FiO2 from 70%to 50 %<br>) |
| Hospital Day 4 | Stop Amiodarone<br>Use Simendan | Maintain CRRT 12 hr.<br>Maintain Meropenem<br>Maintain Thymopentin<br>Maintain Midazolam + Fentanyl<br>Maintain Enteral nutrition<br>Decrease Morphine to 1 mg/h | Invasive Ventilator ( Increase VT from 450 to 500 mL<br>Increase PS from 12 to 15 cm H2O<br>Increase PEEP from 12 t16 cm H2O<br>Decrease Frequency from20 to 18 bpm.<br>) |
| Hospital Day 5 | Stop Morphine | Maintain CRRT 12 hr.<br>Maintain Meropenem<br>Maintain Thymopentin<br>Maintain Midazolam + Fentanyl<br>Increase Enteral nutrition<br>from 1000 to 1500 mL | Intermittent non-invasive ventilato<br>r ( ST mode<br>FiO2 60%<br>IPAP 15 cmH2O<br>EPAP 11 cmH2O<br>Frequency 20 bpm.<br>) |
| Hospital Day 6 | Use Human albumin (40g, 10 mg/h) | Maintain CRRT 12 hr.<br>Maintain Meropenem<br>Maintain Thymopentin<br>Maintain Midazolam + Fentanyl | Invasive Ventilator ( Increase VT from 500 to 550 mL<br>Increase PS from 12 to 15 cm H2O<br>Decrease PEEP from 16 to 11 cmH2O<br>Increase FiO2 from 50% to 70 %<br>Increase Frequency from18 to 24 bpm.<br>) |
| Hospital Day 7 | Stop Meropenem<br>Use Caspofungin + Tezhixing<br>+ Daltotropin<br>Use Amiodarone (oral) | Maintain CRRT 8 hr.<br>Maintain Thymopentin<br>Maintain Midazolam + Fentanyl |  |
| Hospital Day 8 | Stop Xuebijing<br>Stop Mucosolvan | Maintain Amiodarone (oral)<br>Maintain CRRT 8 hr.<br>Maintain Caspofungin + piperacillin<br>+ tazobactam<br>+ Daltotropin<br>Maintain Abidor<br>Maintain Thymopentin<br>Maintain Midazolam + Fentanyl<br>Maintain Human albumin (at least 30g, 10 mg/h)<br>Decrease Enteral nutrition<br>from 1500 to 1000 mL | Invasive Ventilator ( Increase VT from 550 to 650 mL<br>Decrease PEEP from 11 to 10 cmH2O<br>Increase FiO2 from 70% to 100 %<br>) |
| Hospital Day 9 | Use Remifentanil (0.04 ug/Kg-min)<br>Stop CRRT<br>Stop Caspofungin (1 day) | Maintain Midazolam<br>(3 ug/Kg-min)<br>Maintain piperacillin + tazobactam | Invasive Ventilator ( Decrease FiO2 from 100% to 90%<br>) |
|  | Use Saline (100 mL,<br>total <= 2500 mL) | + Daltotropin<br>Maintain Amiodarone (oral)<br>Maintain Abidor<br>Maintain Thymopentin<br>Maintain Fentanyl |  |
| **Hospital Day 10** | Use Ceylon<br>Use Amiodarone<br>(Intravenous injection, 0.15g)<br>Remove subclavian and<br>femoral vein catheters<br>Use Rivaroxaban | Maintain Midazolam<br>(3 ug/Kg-min)<br>Maintain Remifentanyl<br>(0.04 ug/Kg-min)<br>Maintain Caspofungin<br>+ piperacillin<br>+ tazobactam<br>+ Daltotropin<br>Maintain Amiodarone (oral)<br>Maintain Abidor<br>Maintain Thymopentin<br>Maintain Fentanyl | Invasive Ventilator (<br>Decrease VT from 650<br>to 600 mL<br>Increase PEEP from 11 to 12 c<br>mH2O<br>Decrease FiO2 from 90%to 65<br>%<br>)<br>Left subclavian vein puncture cath<br>eterization |
| **Hospital Day 11** | Use Norepinephrine<br>(0.4 ug/Kg-min)<br>Stop Tezhixing<br>Stop Daltotropin<br>Use Supran + Tigecycline<br>Stop Rivaroxaban (10mg) | Maintain Midazolam<br>(3 ug/Kg-min)<br>Maintain Remifentanyl<br>(0.04 ug/Kg-min)<br>Maintain Caspofungin<br>Maintain Amiodarone (oral)<br>Maintain Abidor<br>Maintain Thymopentin<br>Maintain Fentanyl | Invasive Ventilator (<br>Increase FiO2 from 65% to 70<br>%<br>) |
| **Hospital Day 12** | Use Furosemide diuretic<br>+ calcium gluconate<br>Use Trimetazidine tablets<br>+ glycyrrhizin tablets | Maintain Midazolam<br>(3 ug/Kg-min)<br>Maintain Remifentanyl<br>(0.04 ug/Kg-min)<br>Decrease Norepinephrine<br>(from 0.4 to 0.3 ug/Kg-min)<br>Maintain Supran<br>+ Tigecycline<br>+ Caspofungin<br>Maintain Amiodarone (oral)<br>Maintain Abidor<br>Maintain Thymopentin<br>Maintain Fentanyl | Invasive Ventilator (<br>Decrease FiO2 from 70%to 45<br>%<br>)<br>CRRT 6 hr.<br>Left femoral vein catheterization |
| **Hospital Day 13** | Use Bisorol 2.5mg qd. | Decrease Amiodarone<br>to 0.2g qd.<br>Maintain CRRT 8hr.<br>Increase Albumin<br>Decrease Remifentanyl<br>(from 0.04 to 0.01 ug/Kg -min) | Invasive Ventilator (<br>Increase FiO2 from 45% to 55<br>%<br>) |
| **Hospital Day 14** |  | Maintain Midazolam<br>(3 ug/Kg-min)<br>Maintain Remifentanyl<br>(0.01 ug/Kg-min)<br>Maintain Norepinephrine<br>(0.3 ug/Kg-min)<br>Maintain Supran<br>+ Tigecycline<br>+ Caspofungin<br>Maintain Amiodarone (oral)<br>+ Trimetazidine<br>Maintain CRRT<br>Maintain Thymopentin<br>Maintain Fentanyl | Invasive Ventilator (<br>Increase FiO2 from 55% to 60<br>%<br>) |
| **Hospital Day 15** | Stop Supran<br>Use Tezhixing<br>Stop CRRT | Maintain Midazolam<br>(3 ug/Kg-min)<br>Increase Remifentanyl<br>(from 0.01 to 0.04 ug/Kg -min)<br>Maintain Supran<br>+ Tigecycline<br>+ Caspofungin | Blood transfusion (<br>02/16 01:45-03:00<br>A+ 400 mL<br>02/16 20:45-23:00<br>A+ 200 mL<br>02/16 23:25 to<br>02/17 04:00<br>Suspended red blood cells 4U<br>) |
|  |  | Maintain Thymopentin<br>Maintain Fentanyl | Invasive Ventilator ( Decrease PEEP from 12 to 10 cmH2O ) |
| Hospital Day 16 | Stop Anticoagulant related medicine<br>Stop CRRT<br>Use Albumin (20g)<br>Use Somatostatin<br>Use Omeprazole<br>Use Cavin + Vitamins | Maintain Midazolam<br>(3 ug/Kg-min)<br>Maintain Remifentanyl<br>(0.04 ug/Kg-min)<br>Maintain Tezhixing + Tigecycline + Caspofungin | Invasive Ventilator ( Decrease FiO2 from 60%to 50 % ) |
| Hospital Day 17 | Remove femoral vein catheters<br>Stop CRRT | Maintain Cavin (800 mL) + Enteral nutrition (500 mL)<br>Maintain Tezhixing + Tigecycline + Caspofungin | Invasive Ventilator ( Decrease PEEP from 10 to 8 c mH2O ) |
| Hospital Day 18 | Use Propofol (15ug/Kg-min)<br>Stop Somatostatin<br>Stop Omeprazole<br>Use Pantoprazole (oral)<br>Stop Amiodarone<br>Use Bisoral 2.5mg qd.<br>Stop Cavin<br>Stop Tezhixing<br>Use Ceftazidime avitastat<br>Use Albumin (30g)<br>Use Chloroquine (oral) | Decrease Remifentanyl<br>(from 0.04 to 0.02 ug/Kg -min)<br>Maintain Tezhixing + Tigecycline + Caspofungin<br>Increase Enteral nutrition from 500 to 1000 mL | Invasive Ventilator ( Increase PEEP from 8 to9 cmH 2O ) |
| Hospital Day 19 | Use Rivaroxaban<br>Stop Caspofungin<br>Use Voricone<br>Stop Chloroquine phosphate<br>Use Albumin (20g, 10mL/h) | Maintain Tigecycline + Ceftazidime avitastat | Invasive Ventilator ( Decrease FiO2 from 60%to 45 %<br>Decrease VT from 600 to 500 mL<br>Decrease PEEP from 9 to 8 c mH2O<br>Decrease Frequency from24 to 19 bpm ) |
| Hospital Day 20 | Use Linezamide + Caspofungin + Daltotropin<br>Use Somatostatin<br>Use Omeprazole<br>Stop Enteral nutrition<br>Use CRRT 6 hr.<br>Stop Traditional Chinese medicine<br>Use Acetylcysteine nebulization | Decrease Propofol to 20 ug/Kg-min<br>Maintain Midazolam<br>(3 ug/Kg-min)<br>Maintain Remifentanyl<br>(0.04 ug/Kg-min)<br>Maintain Ceftazidime avitastat<br>Increase Ambroxol to 240mg q12h | Invasive Ventilator ( Decrease FiO2 from 45%to 40 %<br>Decrease VT from 500 to 450 mL<br>Decrease PS from 15 to12 cm H2O<br>Decrease PEEP from 8 to 4 c mH2O<br>Increase Frequency from19 to 20 bpm ) |
| Hospital Day 21 | Use Chloroquine (0.4g)<br>Stop Daltotropin<br>Use Vancomycin (0.5g q8h)<br>Use Recovery patient's plasma<br>Use Albumin (20g, 10mL/h)<br>Use CRRT | Maintain Linezamide + Caspofungin + Ceftazidime avitastat | Invasive Ventilator ( Increase FIO2 from 40% to 70 %<br>Decrease VT from 450 to 440 mL<br>Increase Frequency from20 to 35 bpm ) |

**Table S6.**
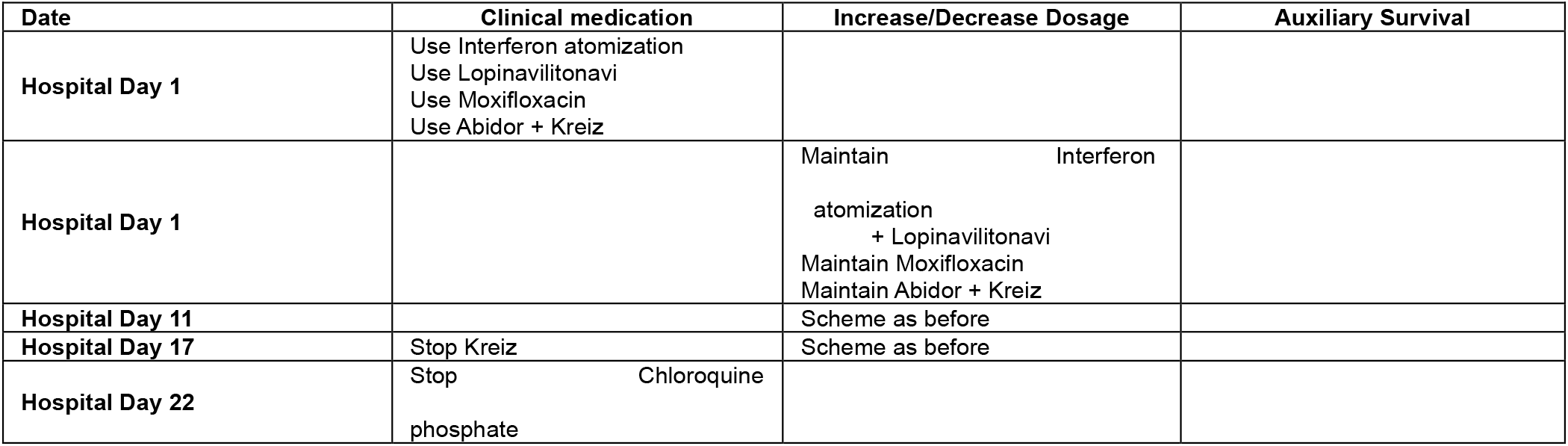
Clinical medication records of the mild case.

**Table S7.** Microbiomes infected in plasma of cases and controls^1^.

| Sample | Species | Coverate rate | Depth | No. of uniquely mapped reads | Type of microbimes |
| --- | --- | --- | --- | --- | --- |
| Control1 | Stenotrophomonas maltophilia | 82.06 | 6.51 | 89132 | Bacteria |
|  | Pseudomonas sp. TKP | 8.31 | 1.08 | 4129 | Bacteria |
|  | Pseudomonas fluorescens | 9.17 | 1.11 | 1088 | Bacteria |
|  | Cuti Bacteriaterium acnes | 28.39 | 1.21 | 7045 | Bacteria |
|  | Elizabethkingia miricola | 8.32 | 1.10 | 1113 | Bacteria |
|  | Elizabethkingia anophelis | 5.38 | 1.09 | 425 | Bacteria |
|  | Staphylococcus capitis | 4.30 | 1.04 | 876 | Bacteria |
|  | Staphylococcus haemolyticus | 3.45 | 1.03 | 851 | Bacteria |
|  | Variovorax paradoxus | 6.10 | 1.17 | 639 | Bacteria |
| Control2 | Stenotrophomonas maltophilia | 78.30 | 4.27 | 56407 | Bacteria |
|  | Pseudomonas sp. TKP | 9.79 | 1.08 | 4937 | Bacteria |
|  | Pseudomonas fluorescens | 12.09 | 1.14 | 1430 | Bacteria |
|  | Cuti Bacteriaterium acnes | 9.07 | 1.06 | 2014 | Bacteria |
|  | Elizabethkingia miricola | 4.45 | 1.06 | 608 | Bacteria |
|  | Variovorax paradoxus | 6.02 | 1.15 | 628 | Bacteria |
| Mild1 | Chryseo Bacteriaterium indologenes | 46.02 | 1.49 | 32259 | Bacteria |
|  | Cuti Bacteriaterium acnes | 7.48 | 1.05 | 1661 | Bacteria |
|  | Methylophilus methylotrophus | 6.43 | 1.05 | 1437 | Bacteria |
| Mild2 | Chryseo Bacteriaterium indologenes | 42.17 | 1.42 | 28162 | Bacteria |
|  | Methylophilus methylotrophus | 5.19 | 1.03 | 1148 | Bacteria |
| Mild3 | Chryseo Bacteriaterium indologenes | 35.68 | 1.33 | 22399 | Bacteria |
|  | Methylophilus methylotrophus | 4.27 | 1.03 | 940 | Bacteria |
| Mild4 | Chryseo Bacteriaterium indologenes | 47.46 | 1.54 | 34338 | Bacteria |
|  | Methylophilus methylotrophus | 7.29 | 1.05 | 1641 | Bacteria |
| Severe3 | Chryseo Bacteriaterium indologenes | 4.99 | 1.03 | 2326 | Bacteria |
| Severe4 | Chryseo Bacteriaterium indologenes | 5.50 | 1.04 | 2658 | Bacteria |
| Severe3 | Human betaherpesvirus 5 | 10.60 | 1.10 | 249 | Virus |
| Severe4 | Human betaherpesvirus 5 | 74.81 | 2.18 | 3593 | Virus |
|  | Human alphaherpesvirus 1 | 3.54 | 1.03 | 52 | Virus |

1. Microbiomes with no. of uniquely mapped reads above 10 and coverage rate (coverage of viruses divided by coverage of autosomes) above 3 were included.

**Table S8.**
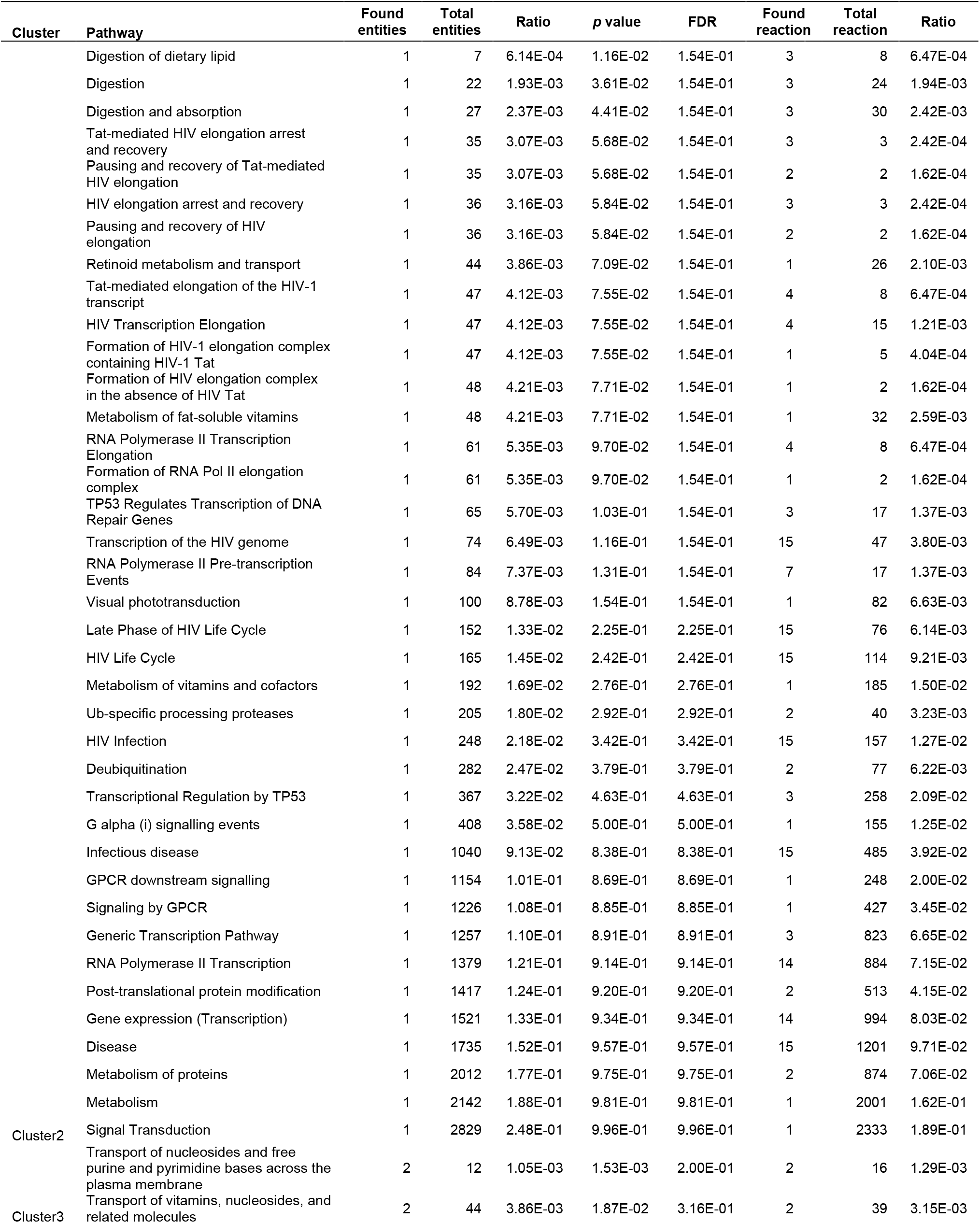
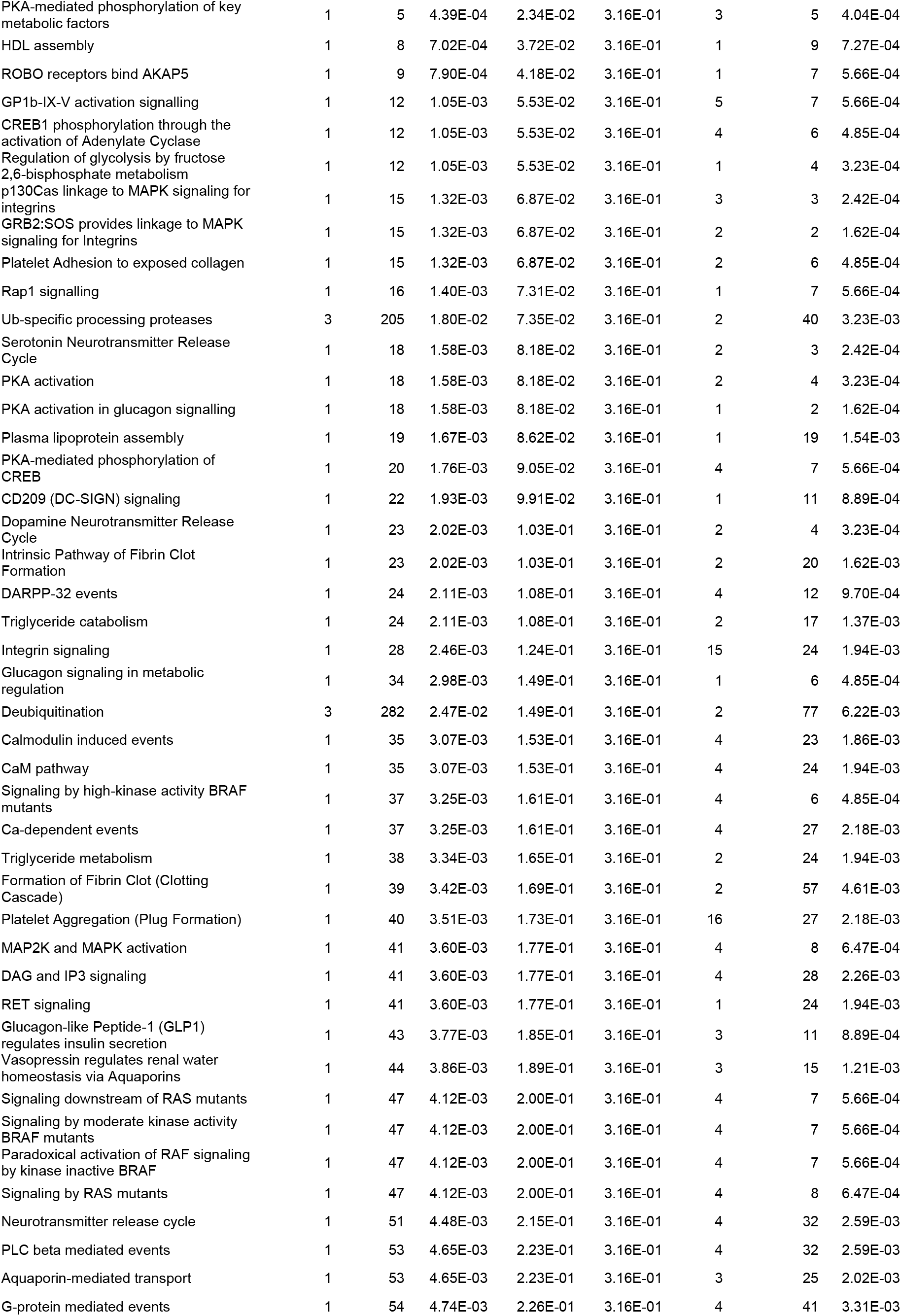
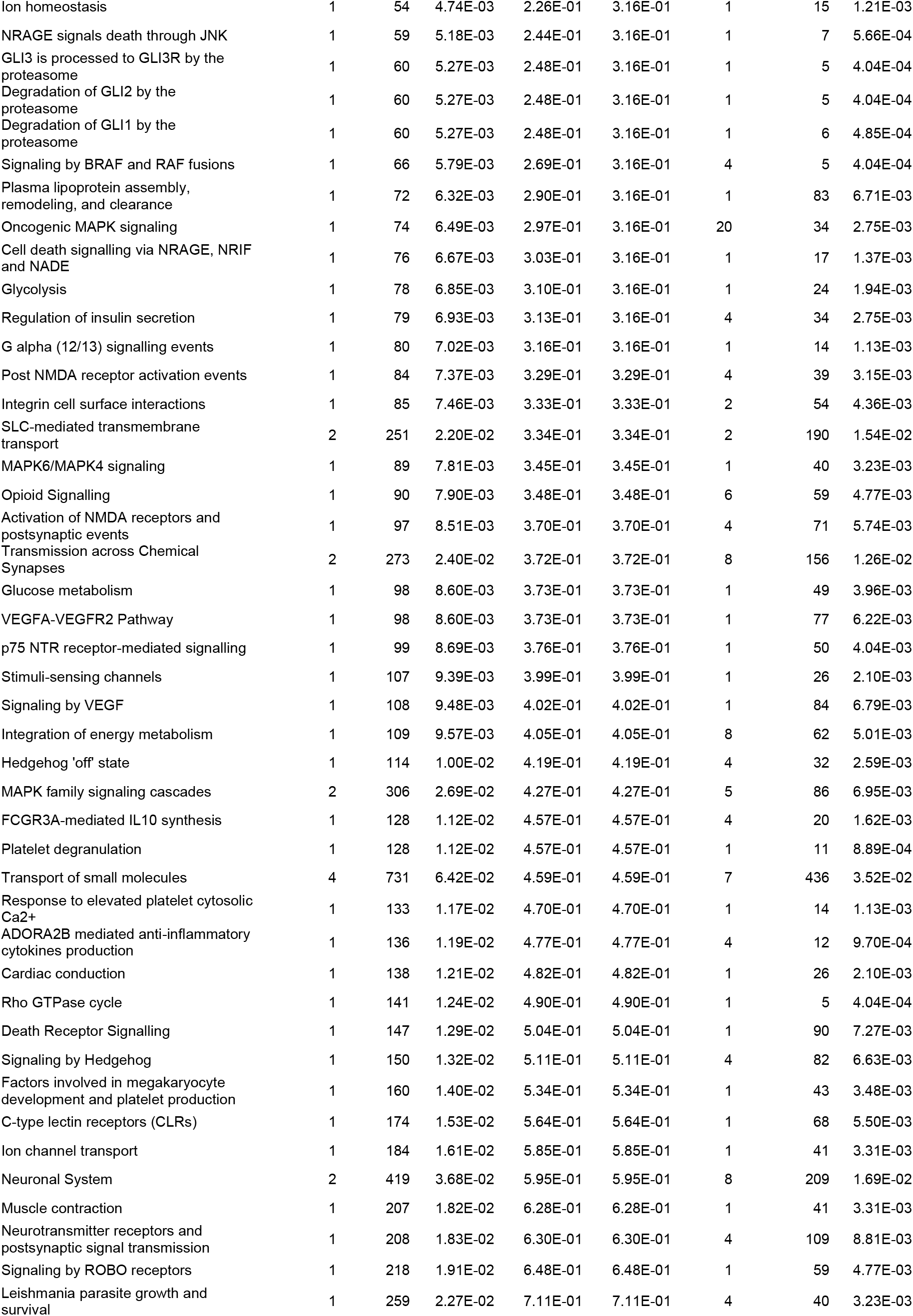
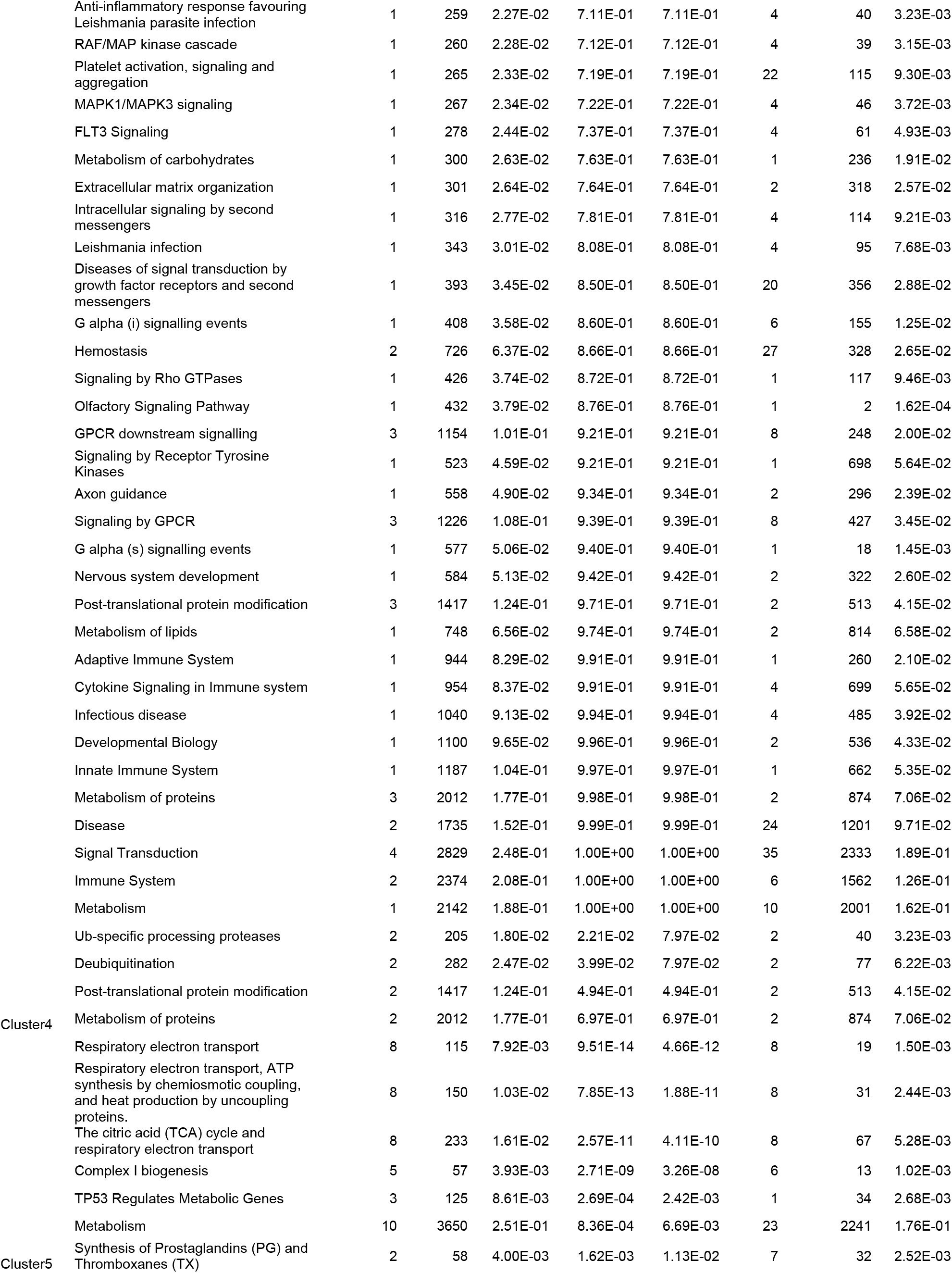
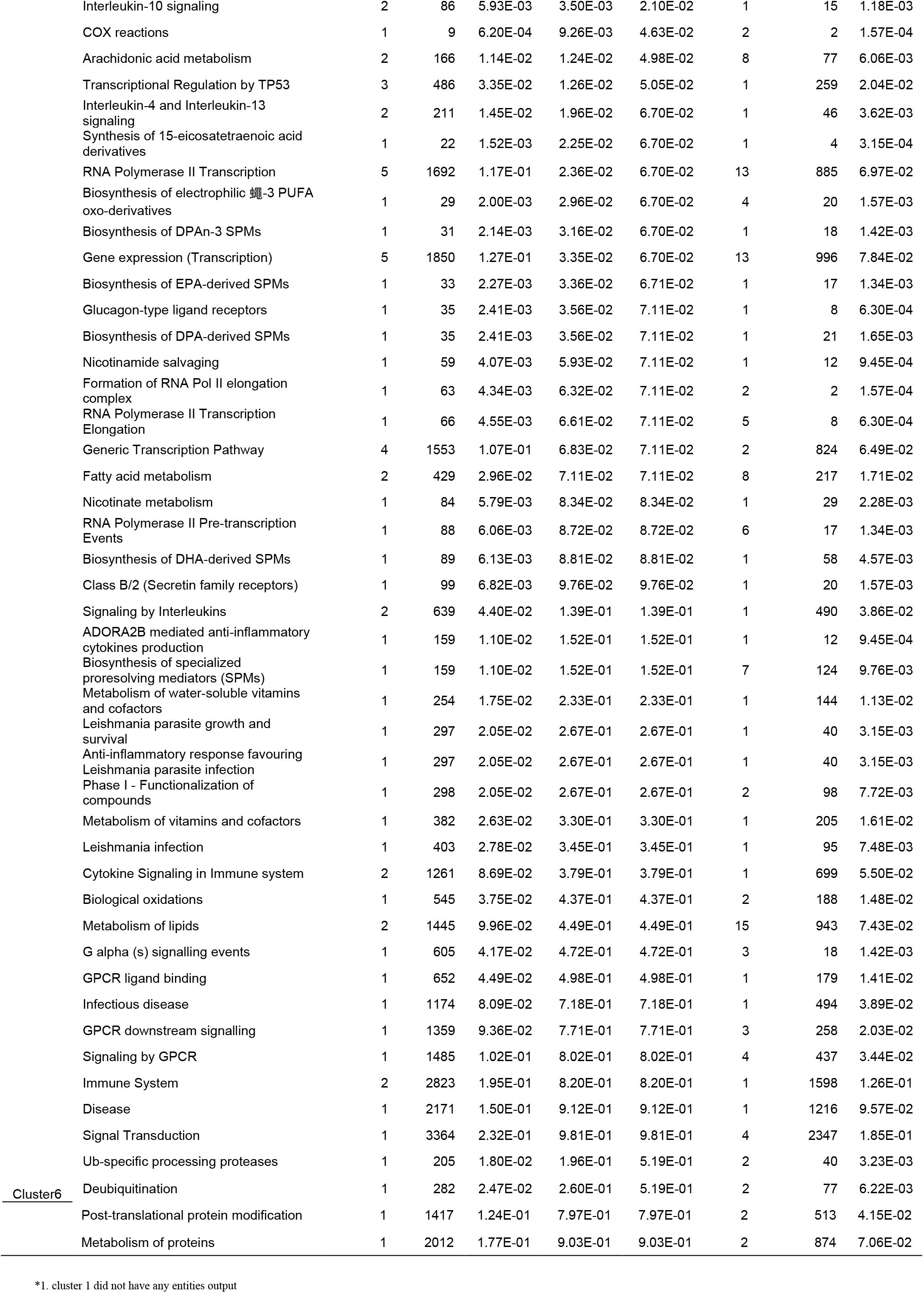
Results on pathway analysis of the 6 clusters from coverage around 200bp-TSSs from cfDNA of one mild case, one severe case, and two controls^1^.

